# Single-cell and spatial profiling of cysteine cathepsins identifies tumor states relevant to antibody-drug conjugates in breast cancer

**DOI:** 10.64898/2026.05.10.26352827

**Authors:** Natalia Ćwilichowska-Puślecka, Natalia Małek-Chudzik, Oliwia Gorzeń, Tobiasz Puślecki, Jakub Mlost, Julia Nguyen, Bartosz Dołęga-Kozierowski, Piotr Kasprzak, Mirosław Sopel, Katarzyna Groborz, Bartłomiej Szynglarewicz, Rafał Matkowski, Marcin Poręba

**Affiliations:** Faculty of Chemistry, Wroclaw University of Science and Technology, 50-370 Wroclaw, Poland; Faculty of Information and Communication Technology, Wroclaw University of Science and Technology, 50-370 Wroclaw, Poland; Science for Life Laboratory, Department of Biochemistry and Biophysics, Stockholm University, SE-106 91 Stockholm, Sweden; Lower Silesian Oncology, Pulmonology and Hematology Center, Wroclaw, Poland; Faculty of Medicine, Wroclaw University of Science and Technology, 50-370 Wroclaw, Poland; SBP Medical Discovery Institute, La Jolla, 92037, CA, USA; Wrocław Medical University, Faculty of Medicine, Department of Oncology, 50-367

## Abstract

Breast cancer is a highly heterogeneous disease shaped by dynamic interactions between malignant cells, immune infiltrates, stromal compartments, and the extracellular matrix. Among the molecular regulators of these interactions, cysteine cathepsins and legumain have emerged as important proteases involved in tissue remodeling, immune regulation, and tumor progression, yet their distribution and functional status across human breast cancer ecosystems remain insufficiently defined. Here, we performed an integrated protease-centric analysis of breast cancer specimens from 66 patients using high-dimensional single-cell mass cytometry of matched peripheral blood and tumor samples, imaging mass cytometry of intact tissues, and activity-based TOF probes for in situ detection of active proteases. Systemic immune profiling identified two patient clusters associated primarily with neoadjuvant therapy and tumor grade, accompanied by distinct cytokine and circulating protease patterns. In tumors, single-cell analysis revealed pronounced interpatient heterogeneity in tissue architecture and immune infiltration, while protease profiling uncovered reproducible cell type-associated modules, including cathepsin B/L-cystatin C and legumain-cystatin E/M axes. Cathepsins B and L were prominent in tumor-infiltrating immune cells and variably expressed in epithelial cells, whereas cathepsin D showed broader tumor distribution and cathepsin S remained more restricted. In epithelial cells, HER2 expression did not consistently coincide with high cathepsin B or L abundance, enabling identification of a limited subgroup of patients with combined HER2-high/protease-high states relevant to protease-cleavable antibody-drug conjugates. Spatial imaging further localized cathepsins B and D to tumor-stroma interfaces and macrophage-rich niches, and activity-based IMC confirmed the presence of catalytically active cathepsin B in human breast tumor tissue. Together, these findings define cysteine cathepsins as spatially and cellularly organized components of breast tumor ecosystems and provide a framework for protease-informed patient stratification and biomarker-protease pairing in targeted therapy.

## Introduction

Breast cancer remains one of the most common malignancies worldwide and represents a highly heterogeneous disease at both the molecular and cellular levels. Clinically, breast cancers are broadly classified into hormone receptor-positive luminal subtypes, HER2-enriched tumors, and triple-negative breast cancer (TNBC), each associated with distinct biological behavior, therapeutic vulnerabilities, and patient outcomes (1, 2). However, even within these categories, substantial inter- and intra-tumoral heterogeneity exists, driven not only by genetic diversity among cancer cells but also by the complex and dynamic tumor microenvironment (TME) (3). The TME comprises immune cells, fibroblasts, endothelial cells, and extracellular matrix components that collectively influence tumor progression, immune evasion, and therapeutic response (4). Increasing evidence suggests that understanding breast cancer requires integrative approaches that resolve cellular diversity and spatial organization within intact tissues rather than relying solely on bulk molecular measurements (3, 5, 6). Mass cytometry (CyTOF) has emerged as a powerful technology for high-dimensional single-cell analysis, enabling the simultaneous quantification of dozens of protein markers per cell using metal-tagged antibodies (7, 8). By overcoming the spectral limitations of fluorescence-based cytometry, CyTOF allows deep phenotyping of heterogeneous tumor samples, including detailed characterization of immune and stromal compartments in breast cancer (9, 10). Application of CyTOF to dissociated breast tumors has revealed extensive diversity among tumor-infiltrating immune cells and identified cellular states associated with prognosis and treatment response (9). However, tissue dissociation inherently disrupts spatial context, which is critical for understanding cell-cell interactions and tissue architecture. Imaging mass cytometry (IMC) addresses this limitation by combining CyTOF detection with laser ablation of metal-labeled tissue sections, enabling highly multiplexed protein imaging at single-cell resolution within intact tissues (11). IMC has been successfully applied to breast cancer to map tumor architecture, stromal niches, and immune infiltration patterns, revealing spatially defined microenvironments that correlate with clinical outcomes (12–14). These studies have demonstrated that spatial relationships-such as immune cell exclusion, compartmentalization of fibroblast subsets, or localized signaling niches-carry prognostic and biological significance beyond cell composition alone. As such, IMC has become a cornerstone technology for spatial tumor biology and patient stratification.

Among the molecular drivers shaping the breast cancer microenvironment, proteases play an important role. Proteolytic enzymes regulate extracellular matrix remodeling, tumor invasion, angiogenesis, immune modulation, and therapy resistance (15, 16). Cysteine cathepsins constitute a prominent family of lysosomal proteases that are frequently dysregulated in cancer (17, 18). In breast cancer, cathepsins such as B, L, S, and D are often overexpressed and mislocalized, exhibiting extracellular and pericellular activity that promotes invasion and metastasis (17, 19). These enzymes degrade structural matrix components, activate growth factors, and process cell surface receptors, thereby reshaping both tumor architecture and signaling landscapes. Importantly, cysteine cathepsins are expressed not only by cancer cells but also by tumor-associated macrophages, fibroblasts, and other stromal populations, underscoring their relevance within the TME. Legumain (asparagine endopeptidase) is another lysosomal cysteine protease increasingly implicated in cancer biology (20, 21). Highly expressed in tumor-associated macrophages and aggressive tumor cells, legumain contributes to extracellular matrix degradation and can activate other proteases, including cysteine cathepsins, amplifying proteolytic cascades within tumors (22). Elevated expression and activity of cathepsins and legumain in breast cancer have been associated with poor prognosis and therapy resistance, making them attractive candidates for both biomarker discovery and therapeutic targeting (17–19, 23). Despite their importance, most studies of proteases in cancer rely on measurements of gene or protein expression, which do not necessarily reflect enzymatic activity. Proteases are tightly regulated at multiple post-translational levels, and only their active forms directly mediate biological effects. To address this gap, chemical approaches such as activity-based probes (ABPs) have been developed to selectively and covalently label active proteases (24–28). ABPs targeting cysteine cathepsins and legumain have enabled visualization of protease activity in cells, tissues, and *in vivo* tumor models, including applications in fluorescence-guided surgery (29–31). However, conventional fluorescent probes are limited in multiplexing capacity, restricting simultaneous analysis of multiple enzymatic activities. Recently, mass cytometry-compatible activity-based probes (TOF-probes) have been introduced to overcome this limitation by labeling active enzymes with distinct lanthanide isotopes (32, 33). These probes enable multiplexed detection of protease activity using CyTOF and IMC, conceptually extending the notion of the proteome to the “activome”. While this strategy has been validated in cell systems, its application to human tumor tissues remains largely unexplored. In this study, we leverage mass cytometry and imaging mass cytometry to perform an in-depth analysis of cysteine cathepsins and legumain in human breast cancer. Using metal-tagged antibody panels, we systematically profile protease expression across tumor, immune, and stromal compartments in both dissociated tumors and intact tissue sections, providing a comprehensive spatial and single-cell view of protease distribution within the breast cancer microenvironment. Furthermore, we present proof-of-concept imaging of cysteine cathepsin activity in breast cancer tissue sections using a TOF-compatible activity-based probe. By integrating expression-based and functional protease imaging within the IMC framework, this work establishes a foundation for studying proteolytic activity directly in human tumor tissues and highlights the potential of combining spatial proteomics with chemical biology to interrogate functional states of the tumor microenvironment.

## Experimental section

### Study design

This study was conducted in accordance with protocols approved by the Bioethical Commission of Wroclaw Medical University (KB-135/2021) and the Institutional Review Board of the Lower Silesian Oncology, Hematology, and Pulmonology Center. Between January 2022 and March 2023, peripheral blood samples and breast tissue specimens were collected from 66 patients with histopathologically confirmed breast cancer who were treated at the Lower Silesian Oncology, Hematology, and Pulmonology Center in Wroclaw, Poland. All patients provided written informed consent prior to enrollment. None of the patients receiving neoadjuvant therapy had been treated with immunotherapy. Exclusion criteria included a positive PCR test for SARS-CoV-2, the presence of other communicable diseases, including hepatitis or HIV, and a tumor size insufficient to obtain a representative sample for testing. Detailed clinical data were obtained from patients’ medical records. Additional clinical information, including smoking status and family history of breast cancer, was obtained through patient questionnaires. Peripheral blood samples were collected one day prior to surgery, before the administration of any preoperative medication. Tissue specimens were collected intraoperatively. Patients represented multiple breast cancer subtypes and received either neoadjuvant therapy (NAT) or adjuvant therapy. Among the study cohort, 34 patients received NAT and 33 received adjuvant therapy. The mean age was 58.0 years in the NAT group and 61.8 years in the adjuvant group. Treatment decisions were based on standard diagnostic procedures, including breast and axillary ultrasound, mammography, breast MRI in selected cases, histopathology (HE, IHC, FISH, ISH), and additional imaging studies only in patients with signs or symptoms of metastatic disease or in clinically high-risk patients. Tumor staging was performed according to the TNM classification system. From a subgroup of 55 patients selected on the basis of tumor size and availability of surrounding tissue, additional samples of tumor tissue and tumor-associated adipose tissue were collected. Samples were processed for formalin-fixed, paraffin-embedded (FFPE) and OCT-based analyses.

### Panel design for protease detection in whole-blood leukocytes

Immunophenotyping of patient-derived whole-blood leukocytes was performed using the Maxpar Direct Immune Profiling Assay (MDIPA, Standard BioTools), supplemented with additional metal-conjugated antibodies targeting selected proteases in unoccupied mass channels. Whole blood was processed by red blood cell lysis prior to staining, and the resulting leukocyte fraction was used for downstream mass cytometry analysis. The following antibodies were included: anti-calpain-1 (^142^Nd, clone A2A), anti-cathepsin B (^165^Ho, clone JA11-02), anti-cathepsin L (^162^Dy, clone 204101), anti-cathepsin S (^175^Lu, clone 1F9), and anti-legumain (^111^Cd, clone 312114). All metal-conjugated antibodies were initially prepared at a concentration of 1 µg/mL, titrated empirically, and used at a final dilution of 1:50.

### Panel design for single-cell breast tumor profiling

Single-cell profiling of breast cancer tumor samples was performed using a custom-designed panel of metal-conjugated antibodies. The panel was composed of antibodies targeting tumor architecture, tumor-infiltrating immune cells, cell death markers, cancer-associated biomarkers, and proteases together with their endogenous inhibitors. Tumor architecture markers included anti-FAP (^164^Dy, clone F11-24), anti-αSMA (^161^Dy, clone 1A4), anti-EpCAM (^141^Pr, clone 9C4), anti-cadherin-3 (^172^Yb, clone 67A4), and anti-CD31 (^144^Nd, clone WM59). Tumor-infiltrating immune cell markers included anti-CD11c (^147^Sm, clone C3.9), anti-CD11b (^209^Bi, clone ICRF44), anti-HLA-DR (^174^Yb, clone L243), anti-CD45 (^89^Y, clone HI30), anti-CD16 (^148^Nd, clone 3G8), anti-CD3 (^154^Sm, clone UCHT1), anti-CD14 (^160^Gd, clone M5E2), anti-CD4 (^110^Cd, clone RPA-T4), anti-CD8 (^114^Cd, clone RPA-T8), anti-CD20 (^171^Yb, clone 2H7), anti-CD15 (^150^Nd, clone W6D3), anti-CD56 (^176^Yb, clone HCD56), anti-CD33 (^167^Er, clone WM53), anti-CD66b (^113^Cd, clone 6/40c), anti-CD19 (^165^Ho, clone FMC63), anti-CD25 (^169^Tm, clone 2A3), and anti-CD44 (^153^Eu, clone OX49). Cell death markers included anti-cleaved PARP (^143^Nd, clone F21-852) and anti-cleaved caspase-3 (^143^Nd, polyclonal). Lysosomal marker was anti-LAMP1 (151Eu, clone H4A3). Cancer-associated proteases and endogenous inhibitors included anti-cathepsin B (^162^Dy, clone 173317), anti-cathepsin L (^158^Gd, clone 204101), anti-cathepsin D (^152^Sm, clone 185111), anti-cathepsin S (^155^Gd, clone 248718), anti-legumain (^173^Yb, clone 312114), anti-cystatin C (^166^Er, clone 197807), anti-cystatin B (^149^Sm, clone 225228), and anti-cystatin E/M (^175^Lu, clone 211515).

### Antibody conjugation

Antibodies were labeled using the Maxpar X8 Antibody Labeling Kit or Maxpar MCP9 Antibody Labeling Kit (Standard Biotools) for lanthanide and cadmium isotopes, respectively, following the manufacturer’s instructions. For lanthanide labeling, 100 μg of antibody was reduced with 4 mM TCEP (Sigma-Aldrich, 51805-45-9) for 30 min at 37°C. The Maxpar X8 polymer was loaded with 2.5 mM lanthanide solution for 1 h, washed with L and C buffers using 3 kDa centrifugal filters (Amicon, Merck, UFC500396), and incubated with the reduced antibody for 1.5 h at 37°C. Conjugates were washed four times with W buffer, concentrated using 50 kDa filters (Amicon, Merck, 505096), and resuspended in antibody stabilizer (Sigma-Aldrich, 55514). Concentrations were determined spectrophotometrically (BioPhotometer D30, Eppendorf) at A280 and adjusted to 0.5 mg/mL. For cadmium labeling, antibodies were processed using the MCP9 polymer, which was pre-washed with L and C buffers. Conjugates were washed in W buffer using 100 kDa centrifugal filters (Amicon, Merck, UFC510096) at 5000 × g for 10 min and stored sealed at 4°C.

### Whole-blood leukocyte staining for mass cytometry

Peripheral blood samples were processed within 2 h of collection. For mass cytometry-based immune profiling, red blood cells were removed by lysis and the resulting leukocyte fraction was used for surface and intracellular staining. Red blood cells were lysed with RBC lysis buffer (BioLegend, 420302). Briefly, 3.5 mL lysis buffer was added to 1 mL of whole blood and incubated for 7 min in the dark, followed by dilution with 10 mL PBS (Sigma-Aldrich, D8537) and centrifugation (400 × g, 5 min). Lysis was repeated for 10 min, and cells were washed three times with Cell Staining Buffer (CSB; Standard Biotools, 201068). Cells were resuspended in CSB, counted using trypan blue exclusion (0.4% trypan blue, Invitrogen, 15250061) on a chamber slide (Invitrogen, 100078809) with an automated counter (Countess 3, Invitrogen). For each staining, 1.2-1.5 × 10^6^ viable cells were used. For immune profiling, cells were blocked with 5% BSA (Gibco, A5256801) in DPBS (Sigma-Aldrich, D8537) for 30 min and stained with the Direct Immune Profiling Assay (DIP; Standard Biotools, S10310) according to the manufacturer’s protocol. Antibodies were resuspended in 300 μL CSB and incubated with cells. After staining, cells were washed with CSB and fixed in 1.6% paraformaldehyde (Thermo Fisher Scientific, 28908) for 10 min, followed by incubation overnight at 4°C in Fix and Perm Buffer (Standard Biotools, S00092) containing 0.1 μM Iridium Intercalator (Standard Biotools, 201192A). For intracellular protease staining, permeabilization was performed with the eBioscience Permeabilization Kit (00-5523-00). Cells were blocked, incubated with antibody cocktails (antibodies centrifuged at 13,000 × g for 2 min before use), washed, fixed, and stored as described above.

### Plasma collection and storage

Peripheral blood (10 mL) was collected into EDTA-coated tubes and diluted 1:1 with phosphate-buffered saline (PBS) containing 2% fetal bovine serum (FBS; Gibco, A5256801). Diluted blood was layered onto Lymphoprep (Serumwerk Bernburg, 1858) and centrifuged at 800 × g for 30 min at room temperature without brake. Plasma was carefully collected, aliquoted (1 mL), snap-frozen in liquid nitrogen, and stored at −80°C until further analysis.

### Cytokine measurement

Plasma cytokine levels were quantified using the Luminex xMAP technology on a Luminex 200 System (Luminex Corporation, Austin, TX) with the MILLIPLEX Human High Sensitivity T Cell Magnetic Panel (Merck, HSTCMAG-28). Plasma samples were thawed on ice, vortexed, and centrifuged at 1,000 × g for 5 min to remove debris. Twenty-five microliters of standards, controls, or plasma samples were incubated with premixed magnetic beads overnight at 4°C with continuous shaking. Plates were washed and incubated sequentially with detection antibodies (25 μL, 30 min) and streptavidin-phycoerythrin (25 μL, 30 min). After final washes, beads were resuspended in Sheath Fluid Plus and acquired on the Luminex 200 system, recording a minimum of 50 beads per analyte. Data were expressed as median fluorescence intensity (MFI) and analyzed using Belysa Immunoassay Curve Fitting Software. The analytes included IL-1β, IL-6, TNFα, IL-10, IL-12, IFNγ, IL-21, IL-4, IL-5, IL-13, IL-17A, IL-23, MIP-1α, MIP-1β, MIP-3α, IL-8, I-TAC, fractalkine, GM-CSF, and IL-7.

### Breast cancer tissue collection, storage, and dissociation

Fresh breast cancer specimens, including tumor tissue and adjacent cancer-associated adipocytes (CAAs), were collected by pathologist and surgeon in MACS Tissue Storage Solution (Miltenyi Biotec, 130-100-008) and maintained at 4°C. Tumor tissues were processed within 24 h of collection, whereas CAAs were snap-frozen and stored at −80°C. Tissue dissociation was performed using the Human Tumor Dissociation Kit (Miltenyi Biotec, 130-128-030) on a gentleMACS OctoDissociator with heating, following the manufacturer’s breast cancer-specific protocol. After dissociation, samples were filtered through 70 μm strainers (Miltenyi Biotec, 130-098-463) to remove residual aggregates. Cell viability was assessed by trypan blue exclusion, and only viable single-cell suspensions of sufficient quality were used for downstream staining and acquisition. Samples with extensive debris, poor viability, or insufficient cell yield were excluded from further analysis. For tumor architecture analyses, only samples yielding more than 10,000 acquired cells were retained, as indicated in the Results section.

### FFPE and OCT tissue processing

For formalin-fixed paraffin-embedded (FFPE) samples, tissues were fixed in 4% buffered formaldehyde (pH 6.9; Sigma-Aldrich, 100496), paraffin-embedded, and sectioned at a thickness of 10 μm. For cryopreservation, tumor samples were embedded in optimal cutting temperature (OCT) compound (Leica, 14020108926), snap-frozen at −80°C, and sectioned at −18 to −16°C (10 μm) using a cryostat (Leica CM1860 UV). Tissue sections were mounted onto SuperFrost Plus slides (Fisher Scientific, 12-550-15) and stored at −80°C until staining.

### Staining of dissociated tumor cells

Single-cell suspensions obtained from dissociated tumor tissues were adjusted to the desired concentration and blocked in DPBS containing 5% bovine serum albumin for 45 min at room temperature. Cells were then incubated with surface antibody cocktails for 45 min at room temperature. After washing with CyTOF staining buffer, cells were fixed and permeabilized using the eBioscience Intracellular Fixation and Permeabilization Kit according to the manufacturer’s instructions. Intracellular antibody staining was subsequently performed in CyTOF staining buffer for 30-45 min. Cells were then fixed in 1.6% paraformaldehyde and incubated overnight at 4°C in Fix and Perm Buffer containing 0.1 μM Iridium DNA Intercalator. Prior to acquisition, cells were washed in Cell Staining Buffer and Cell Acquisition Solution and resuspended in acquisition solution containing EQ Four Element Calibration Beads. Tumor samples were processed and acquired individually rather than by barcoding.

### Staining of FFPE sections

FFPE tissue sections were dewaxed in xylene, rehydrated through graded ethanol solutions (96%, 95%, 80%, and 70%), and rinsed in deionized water. Antigen retrieval was performed in high-pH retrieval buffer (Invitrogen, 00-4956-58) at 96°C for 30 min using a Dako Link PT system (Agilent, 20027). Sections were blocked with 5% BSA and incubated overnight at 4°C with antibody cocktails. After washing, sections were counterstained with 0.1 μM Iridium DNA Intercalator and air-dried overnight prior to imaging. The FFPE IMC panel included antibodies against α-SMA, pan-keratin, vimentin, collagen I, CD4, CD8, CD20, CD68, cathepsin B, cathepsin S, and cathepsin D, together with DNA intercalator.

### OCT-embedded tissue and TOF-probe staining

After retrieval from −80°C storage, OCT sections were equilibrated to room temperature, washed with DPBS, fixed in 4% formaldehyde for 30 min, and permeabilized with 0.05% Tween-20 in PBS for 30 min. Following an additional PBS wash, sections were blocked with SuperBlock for 30 min in a humidified chamber. Sections were washed in cathepsin assay buffer (100 mM NaCl, 100 mM sodium acetate, and 10 mM DTT at pH 5.5) and, where indicated, preincubated with the pan-cathepsin inhibitor E64 (25 μM) for 1 h at 37°C prior to probe addition. Control and inhibitor-treated stainings were performed on adjacent serial sections of the same tissue specimen. TOF-compatible activity-based probes were diluted in cathepsin buffer to a final concentration of 1 μM and applied to the sections for 1 h at 37°C. After incubation, slides were washed with PBS, counterstained with 0.125 μM DNA intercalator for 30 min, washed twice with distilled water, air-dried overnight, and stored in a sealed container with silica gel desiccant until acquisition. Probe concentration was selected on the basis of preliminary optimization, as higher concentrations increased background staining in OCT sections.

### Cathepsin B TOF-probe

TOF-probes used in this study belong to a class of mass cytometry-compatible activity-based probes composed of three elements: a protease-selective peptide recognition sequence, an electrophilic warhead that covalently binds the catalytic site of the active enzyme, and a DOTA-chelated stable metal isotope for CyTOF/IMC detection. The cathepsin B-selective probe used for tissue imaging, TOF-120, contained the sequence DOTA(Lu)-PEG(4)-Cha-Leu-hSer(Bzl)-Arg-AOMK. Probe sequences were designed on the basis of HyCoSuL substrate specificity profiling and validated against recombinant proteases prior to tissue application (32, 34). In the current manuscript, TOF-120 was used at 1 μM for OCT tissue labeling and its specificity was functionally confirmed by loss of signal after preincubation of adjacent sections with the pan-cathepsin inhibitor E64.

### Data acquisition, preprocessing, and statistical analysis

Single-cell samples were acquired on a CyTOF Mass Cytometer (Standard BioTools, Canada). Prior to acquisition, cells were washed with Cell Staining Buffer (CSB) and Cell Acquisition Solution (CAS), resuspended in Cell Acquisition Solution containing EQ Four Element Calibration Beads, filtered through 40 μm strainers, and acquired at a rate below 500 events/s. Instrument performance was monitored daily and signal drift during acquisition was corrected using bead-based normalization. This approach uses the signal of embedded metal standards to generate a time-dependent correction function for each event and thereby minimizes short- and long-term fluctuations in detector sensitivity. After acquisition, bead-positive events, debris, and cell-bead doublets were excluded. Putative coincident events and doublets were removed on the basis of DNA intercalator signal and event length, as prolonged ion-cloud pulses in mass cytometry indicate non-single-cell events. Downstream analysis was performed on viable singlet cells only. Marker intensities were exported as .fcs files and analyzed using Cytobank. viSNE maps were generated using the t-SNE implementation available in Cytobank, and FlowSOM was used where indicated for high-dimensional immune cell phenotyping. For visualization and comparative analyses, transformed marker values or population frequencies were used as specified in the figure legends. For patient-level analyses, immune and tumor cell frequencies were calculated as percentages of the respective parent populations or as numbers of events normalized to the total number of acquired cells, as appropriate. Principal component analysis (PCA) was performed after batch correction using linear regression, and unsupervised clustering was applied using k-means clustering to stratify patients into groups with related immune or protease profiles. Hierarchical clustering was performed using median marker expression values across cell populations. Correlation analyses were performed using Spearman’s rank correlation coefficient. For selected analyses, cell counts were log1p-transformed and converted to z-scores to generate immune-axis metrics. Unless otherwise stated, statistical significance between two groups was assessed using two-sided Welch’s t-test. Data analysis and visualization were performed using Cytobank, GraphPad Prism 10, and BioRender. Raw imaging data were processed using the steinbock pipeline (https://bodenmillergroup.github.io/steinbock/latest/cli/intro/).

### Hyperion image acquisition and IMC data processing

Imaging mass cytometry was performed on a Helios time-of-flight mass cytometer coupled to a Hyperion Imaging System (Standard BioTools, Canada). Regions of interest of approximately 1 mm^2^ were selected for ablation, typically at the tumor margin, and scanned at a spatial resolution of approximately 1 μm using a laser frequency of 200 Hz. Daily instrument calibration was performed using a tuning slide spiked with five metal elements. Raw data were collected using the Standard BioTools CyTOF software and processed using the open-source steinbock pipeline. Raw IMC files were converted into multichannel images, including hot-pixel filtering, followed by cell segmentation and single-cell feature extraction. Segmentation was performed using steinbock, and for each image mean signal intensity per marker and per cell, cell morphology features, and neighborhood information were extracted. We used steinbock as the primary image-processing environment for downstream single-cell and spatial analyses. Representative images were selected on the basis of tissue quality, marker preservation, and architectural interpretability.

### IMC region selection, segmentation, and image analysis

For imaging mass cytometry, regions of interest were selected at the tumor margin, where malignant cells interface with stromal and immune compartments. ROIs were chosen on the basis of tissue morphology and marker preservation, and representative regions were selected for acquisition. Raw IMC data were processed using the steinbock pipeline to generate multichannel images, segment individual cells, and extract single-cell marker intensities and morphological features. Segmentation-derived single-cell data were subsequently used for spatial interpretation and marker co-localization analyses. Representative images shown in the figures were selected on the basis of tissue quality, signal-to-background ratio, and their ability to illustrate the dominant spatial patterns observed in the analyzed samples.

### Detection of cathepsins and cancer biomarkers in breast cancer cell lines

BT-474 (ATCC HTB-20, RRID: CVCL_0179), MCF-7 (ATCC HTB-22, RRID: CVCL_0031), and MDA-MB-231 (ATCC HTB-26, RRID: CVCL_0062) breast cancer cell lines were cultured at 37°C in a humidified incubator with 5% CO_2_. Cells were maintained in Dulbecco’s Modified Eagle Medium (DMEM; Gibco, Cat. No. 41965-039) supplemented with 10% fetal bovine serum (FBS; Gibco, Cat. No. A5256801), L-glutamine, sodium pyruvate, penicillin (100 U/mL), and streptomycin (0.1 mg/mL). BT-474, MCF-7, and MDA-MB-231 cells were harvested, counted to 1 × 10^6^ cells, and resuspended in CSB. Staining was performed in the same way as for the blood samples described above. Cells were first stained with a cocktail of metal-tagged antibodies against membrane-bound markers, including EGFR, HER2, and TROP-2. After fixation and permeabilization, intracellular proteins, including cathepsins B, L, S, and D, legumain, and LAMP-1, were stained. Following ^191/193^Ir intercalation, cells were acquired on a CyTOF instrument, and the data were analyzed to determine protein expression levels in each breast cancer cell line. Data were normalized to facilitate comparison of marker expression across the three cell lines.

### ADCs efficacy in breast cancer cell lines (MTS assay)

Cell viability was assessed using the MTS assay (Promega, CellTiter 96® AQueous One Solution Cell Proliferation Assay, Cat. No. G358B) according to the manufacturer’s protocol. Cells were seeded in 96-well plates at a density of 20,000 cells per well for BT-474 and 10,000 cells per well for MCF-7 and MDA-MB-231. Antibody-drug conjugates containing either a pan-cathepsin-cleavable dipeptide linker (Val-Cit), a cathepsin B-selective peptide linker (CatB ADC), or a cathepsin L-selective peptide linker (CatL ADC), as well as trastuzumab (RRID: AB_3112050) and free MMAE, were applied at a final concentration corresponding to 1 nM MMAE and incubated with the cells for 4 days. As a control for complete cytotoxicity, Triton X-100 (Sigma-Aldrich, Cat. No. X100-500ML) was added to control wells to a final concentration of 1%, followed by addition of MTS reagent and incubation at 37°C for 3 h. Blank wells were included, and nonspecific absorbance was subtracted from the final readings. Cell viability was calculated as a percentage relative to untreated cells. Experiments were performed in triplicate and are presented as mean ± standard deviation.

### Protease inhibition assay

Cathepsins B, L, D, and S were kindly provided by Prof. Boris Turk, whereas legumain, MMP-2, MMP-9, and HNE were purchased from commercial suppliers. Enzyme activity was measured in the appropriate assay buffers using the corresponding optimal fluorogenic substrates, as described in our previous publications (34, 35). Proteases were preincubated in assay buffer for 15 min at 37°C, followed by addition of the TOF-120 probe (5 μM, 15 min). The appropriate fluorogenic substrate was then added, and residual enzymatic activity was measured. Enzyme activity in the absence of inhibitor served as the control to which inhibition was normalized.

## Results and discussion

### Study design and clinical characteristic of breast cancer patients

We designed an integrated workflow to quantify systemic immune states, intratumoral cellular architecture, and protease landscapes in breast cancer at single-cell resolution, and to validate protease functionality using activity-based chemistry (**Fig. 1A**). Pre-operative whole blood was analyzed by mass cytometry and paired plasma was profiled for soluble mediators. In parallel, freshly resected tumors were dissociated for CyTOF-based mapping of major tumor compartments and protease/cystatin proteins, while tissue sections were processed for multiplex IMC antibody imaging and, in a subset of samples, TOF-probe-based activity imaging of cathepsin B. Clinical annotation was integrated downstream to relate immune/protease features to subtype, grade, and neoadjuvant treatment status. The clinical cohort included 66 patients with breast cancer representing a broad spectrum of molecular and clinicopathological characteristics (**Fig. 1B**). The mean age was 60.0 ± 13.4, and most patients were postmenopausal (47/66, 71.2%). The largest molecular subgroup was luminal B-like (HER2-) (27/66, 40.9%), followed by luminal B-HER2+ (19/66, 28.8%), luminal A-like (10/66, 15.2%), triple-negative breast cancer (7/66, 10.6%), and HER2+ non-luminal disease (3/66, 4.5%). Most tumors were hormone receptor-positive, with ER positivity observed in 55/66 patients (83.3%) and PR positivity in 44/66 patients (66.7%), while HER2 amplification assessed by FISH was present in 13/66 cases (19.7%). Consistent with this distribution, the mean ER expression was 70.9 ± 41.5%, the mean PR expression was 44.1 ± 43.9%, and the mean Ki67 index was 32.0 ± 23.6%. With respect to tumor burden, T2 lesions were the most frequent category (33/66, 50.0%), whereas distant metastases were uncommon (3/66, 4.5%). Approximately half of the patients had received neoadjuvant therapy before surgery (32/66, 48.5%).

**Figure 1.**
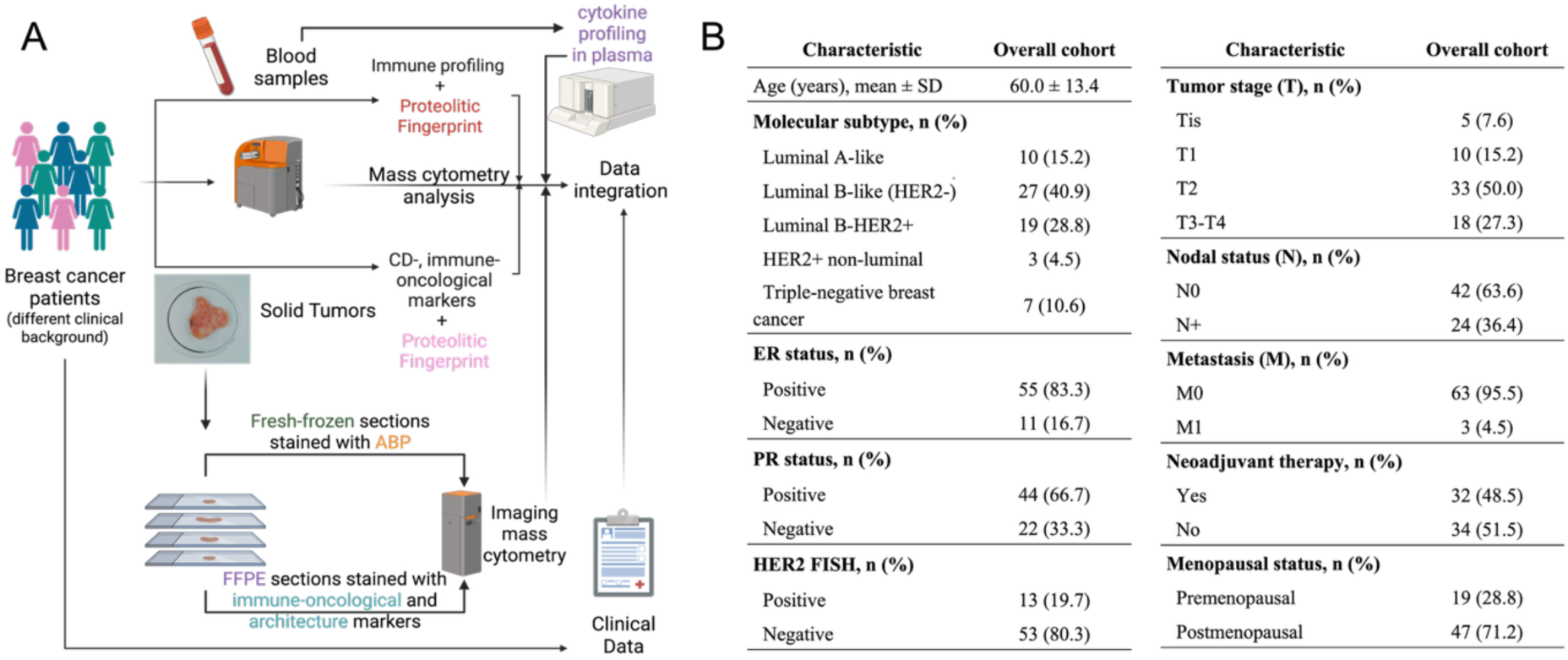
Study design and clinical characteristics of patients. **A** This study aimed to characterize immune, protease, and structural features of breast cancer across clinically diverse patients. Peripheral blood was collected before surgery for cytokine profiling and whole-blood mass cytometry. Tumor samples were dissociated and analyzed with antibody panels to define both immune composition and tissue architecture. Additional tumor material was prepared as FFPE and OCT sections for antibody-based staining and detection of active proteases using TOF-compatible probes. Blood and tumor samples were examined by mass cytometry, including CyTOF and Hyperion imaging, and the resulting datasets were integrated with clinical information for downstream analysis. **B** Clinical characteristics of breast cancer patients analyzed in this study.

### Peripheral blood immune profiling

We first asked whether systemic immune composition in pre-operative blood captured clinically meaningful variation. To address this we performed immune profiling of peripheral blood samples, collected from 66 patients with breast cancer, by mass cytometry (**Fig. 2A**). To visualize the complexity of the peripheral immune compartment, t-SNE analysis was performed using the MDIPA panel. A representative map is shown in **Fig. 2B**, illustrating the distribution of 30 blood cell subtypes identified in peripheral blood. This analysis provided a high-dimensional overview of the systemic immune landscape and served as the basis for subsequent comparisons across clinical subgroups. Next, clinical and blood immune profiling data were integrated and analyzed by principal component analysis (PCA) (**Fig. 2C**). This analysis revealed two major patient clusters, referred to as cluster 1 and cluster 2. When neoadjuvant therapy status (**Fig. 2D**) and nodal status (**Fig. 2E**) were overlaid onto the PCA plot, both variables showed a clear association with cluster distribution. Cluster 1 was enriched in patients who had received neoadjuvant therapy and in patients with higher nodal status, whereas cluster 2 consisted predominantly of patients without neoadjuvant treatment and with lower N stage. Neoadjuvant treatment in this cohort was based mainly on anthracycline- and taxane-containing regimens, including both standard and dose-dense schedules, as well as combinations incorporating cyclophosphamide, epirubicin, paclitaxel, or docetaxel. To further compare the two patient groups, immune cells were aggregated and visualized using viSNE analysis (**Fig. 2F**). At the global level, the maps of cluster 1 and cluster 2 appeared highly similar. However, more detailed quantitative analysis at the patient level revealed clear differences in the abundance of specific immune populations. For each patient, the numbers of monocytes, CD8+ T cells, CD4+ T cells, specialized T cells, NK cells, dendritic cells (DCs), and B cells were calculated per 100,000 blood cells (**Fig. 2G**). This analysis showed that patients in cluster 1 had a lower lymphocyte-to-monocyte ratio and a lower late-to-early NK cell ratio than patients in cluster 2 (**Fig. 2H**). By contrast, the CD4+/CD8+ ratio, the CD8+ effector memory/Treg ratio, and the NK/Treg ratio were comparable between the two clusters (**Fig. S1**). Analysis of individual immune populations further demonstrated that patients in cluster 1 had lower levels of B cells and higher levels of dendritic cells, CD8+ T cells, and monocytes than patients in cluster 2. In contrast, differences in NK cells, specialized T cells, and CD4+ T cells did not reach statistical significance (**Fig. 2I** and **Fig. S2**). A more detailed breakdown of immune cell subtypes is shown in **Fig. S2** and indicates that, for the major populations differing between clusters, similar trends were generally preserved across their respective subpopulations. We next analyzed the data according to breast cancer subtype. In contrast to the cluster-based stratification, subtype-based comparisons revealed a more limited effect on systemic immune composition. Most breast cancer subtypes showed broadly similar immune profiles, whereas the clearest deviations were observed in the TNBC group. Specifically, patients with TNBC displayed higher numbers of specialized T cells and MAIT/NKT cells and lower monocyte counts than the remaining subtypes (**Fig. 2J** and **Fig. S3**). Interestingly, viSNE maps generated for whole-blood immune cells across different breast cancer subtypes, treatment groups, tumor grade categories, or nodal status were broadly similar, indicating that this type of visualization alone was not sufficient to capture the differences observed by quantitative cell population analysis (**Fig. S4**). Together, these findings suggest that breast cancer subtype had only a limited impact on the peripheral immune landscape in this cohort, whereas the cluster-based organization of patients captured more pronounced systemic immune differences. Finally, we examined the relationship between selected immune cell populations and tumor Ki67 status. Correlation analyses were performed for B cells, CD4+ T cells, CD8+ T cells, and monocytes. The B-cell immune axis, defined as the z-score of log1p-transformed B-cell counts, showed a decreasing trend with increasing Ki67 values in patients from cluster 1, whereas no clear trend was observed in cluster 2 (**Fig. 2K**). The remaining immune cell populations did not show a significant association with Ki67 status (**Fig. S5**).

**Figure 2.**
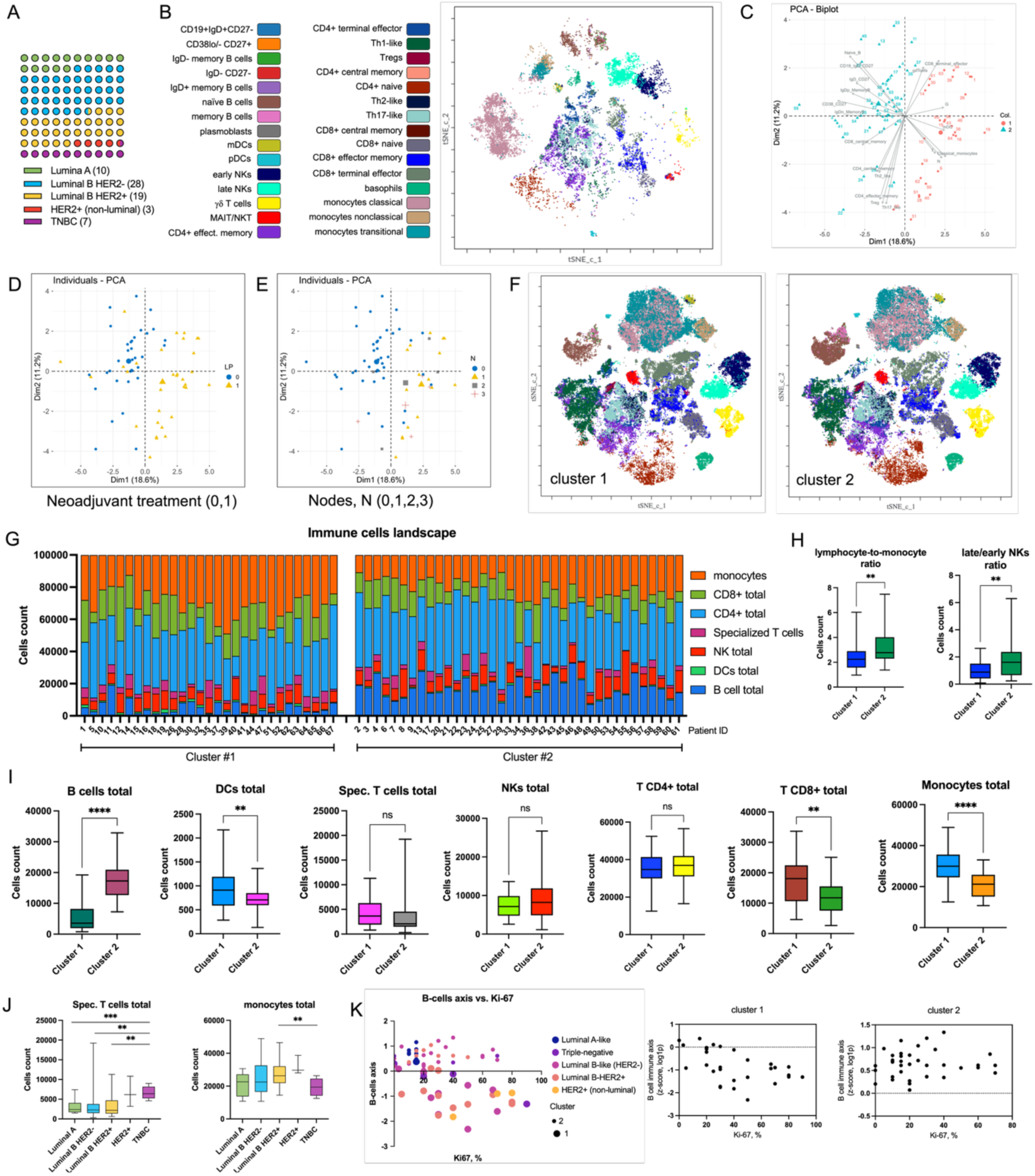
Immune profiling of peripheral blood from breast cancer patients. **A** Distribution of patients stratified by molecular tumor subtype. **B** Representative viSNE analysis of peripheral blood immune cells illustrating the systemic immune landscape. **C** Principal component analysis (PCA) based on immune cell frequencies segregated patients into two distinct clusters. **D** Cluster 1 predominantly included patients who had received neoadjuvant therapy prior to tumor resection. **E** Cluster 1 also showed a strong association with more advanced nodal status. **F** Patients from each cluster were visualized using two independent aggregated viSNE maps, revealing overall similar global immune architectures. **G** Quantitative analysis identified statistically significant differences in the abundance of selected immune cell subsets (measured as single-cell events) between clusters. **H** Statistically significant differences in immune cell ratios, including lymphocyte-to-monocyte and late/early NK cell ratios, between cluster 1 and cluster 2. **I** Differences in major immune cell populations in whole blood between patients assigned to cluster 1 and cluster 2. **J** Differences in specialized T cell counts and total monocyte counts between patients stratified by molecular subtype: Luminal A, Luminal B HER2-, Luminal B HER2+, HER2+ non-luminal, and triple-negative breast cancer (TNBC). **K** Relationship between B cells and tumor Ki67 status. The immune axis (y), defined as the z-score of log1p-transformed cell counts, is plotted against the tumor Ki67 index (0-100%). Data are shown for the entire cohort and stratified by clusters. Statistical significance for all analyses was assessed using Welch’s t-test. * P < 0.05, ** P < 0.005, *** P < 0.0005.

### Protease and cytokine profiling in peripheral blood

We next examined the expression of selected proteases in peripheral blood immune cells and the plasma levels of cytokines and interleukins. The analyzed proteases included cathepsin B, cathepsin L, cathepsin S, cathepsin D, and calpain-1. Cathepsins and calpains are proteolytic enzymes implicated in multiple aspects of breast cancer progression, including tumor invasion, extracellular matrix remodeling, immune regulation, antigen processing and drug resistance (18, 22, 36). Because these processes involve both malignant and immune compartments, profiling protease expression in circulating immune cells may provide insight into systemic alterations associated with breast cancer. To obtain a global overview, viSNE analysis was first performed on aggregated immune cells from patients assigned to cluster 1 and cluster 2 (**Fig. 3A**). We then overlaid the expression of individual proteases onto these maps (**Fig. 3B**). As expected, cathepsin expression was most prominent in monocyte populations, including classical, non-classical, and transitional monocytes, whereas lower expression levels were observed in the remaining immune cell compartments. A more detailed hierarchical clustering analysis across all patients further showed that, in addition to monocytes, cathepsins were also enriched in myeloid dendritic cells, plasmacytoid dendritic cells, and plasmablasts (**Fig. 3C**, **Fig. S6**), which is consistent with the known involvement of these cells in antigen processing, endolysosomal activity, and immune regulation (37). Calpain-1 showed a broadly similar distribution pattern, with the highest expression detected in monocytes, plasmablasts, and dendritic cells, and the lowest levels observed in CD4+ and CD8+ T cells. This pattern was maintained across the entire patient cohort. Importantly, the median expression of proteases also differed between patient clusters. Cathepsin B showed lower expression in several B-cell and T-cell subsets in cluster 2, which largely overlapped with patients who had not received neoadjuvant therapy and presented with lower nodal status (**Fig. 3D**, **Fig. S7A**). Similarly, calpain-1 expression was reduced in multiple B-cell subsets (including naïve CD19+IgD+CD27- and IgD-CD27-cells), T-cell populations (CD4+ naïve and CD8+ naïve), and basophils in cluster 2. Cathepsin L and cathepsin D were also decreased in naïve and memory B-cell subsets, whereas cathepsin S showed lower expression in both IgD+ and IgD-memory B cells in cluster 2 (**Fig. S7B-E**). Notably, none of the analyzed proteases showed consistently higher expression in any immune cell population in cluster 1. These observations suggest that neoadjuvant therapy or mode advanced nodal status may be associated with increased expression of selected proteases in peripheral immune cells. Together, these data indicate that the expression of the analyzed proteases in peripheral blood immune cells was primarily determined by cell identity and was also modulated by cluster assignment or, indirectly, by breast cancer-associated systemic stratification.

**Figure 3.**
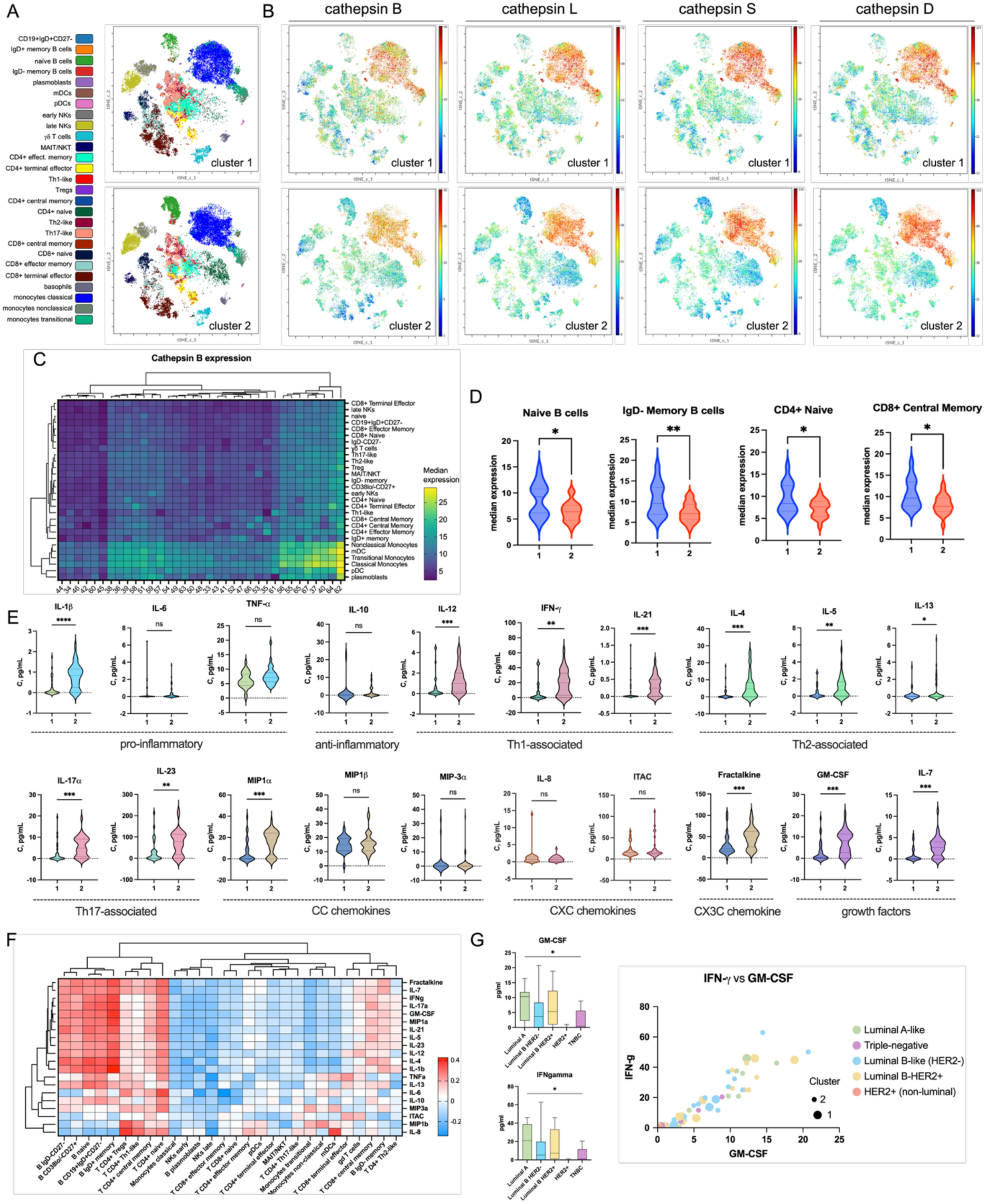
Protease expression and cytokine profiling in peripheral blood of breast cancer patients. **A** viSNE maps of aggregated peripheral blood immune cells from patients assigned to cluster 1 and cluster 2. **B** Overlay of protease expression (cathepsin B, cathepsin L, cathepsin S, cathepsin D, and calpain-1) on viSNE maps. **C** Hierarchical clustering of median expression of cathepsin B across immune cell populations, highlighting enrichment in monocytes, dendritic cells, and plasmablasts. **D** Differences in median expression of cathepsin B between patient clusters, with reduced expression in multiple B-cell and T-cell subsets in cluster 2. **E** Plasma concentrations of cytokines, interleukins, chemokines, and growth factors measured by Luminex, presented as box plots comparing clusters. Cluster 2 shows elevated levels of pro-inflammatory, Th1-, Th2-, and Th17-associated cytokines, as well as chemokines and growth factors. **F** Hierarchical correlation analysis between soluble mediators and immune cell populations, showing positive associations between elevated cytokines and B-cell subsets, as well as CD4+ T-cell populations **G** Cytokine levels stratified by molecular subtype, showing limited differences, with significantly lower GM-CSF and IFNγ levels in TNBC compared with luminal subtypes. Statistical significance for all analyses was assessed using Welch’s t-test. * P < 0.05, ** P < 0.005, *** P < 0.0005.

In the next step, we shifted our focus from cellular protease expression to circulating soluble mediators and analyzed a panel of cytokines, interleukins, chemokines, and growth factors in plasma. The panel included pro-inflammatory cytokines (IL-1β, IL-6, TNFα), an anti-inflammatory cytokine (IL-10), Th1-associated cytokines (IL-12, IFNγ, IL-21), Th2-associated cytokines (IL-4, IL-5, IL-13), Th17-associated cytokines (IL-17A, IL-23), CC chemokines (MIP-1α, MIP-1β, MIP-3α), CXC chemokines (IL-8, I-TAC), the CX3C chemokine fractalkine, and growth factors (GM-CSF and IL-7) (38). Comparison of median concentrations and distribution patterns between the two clusters showed that cluster 2, which was associated predominantly with the absence of neoadjuvant treatment, displayed elevated levels of IL-1β, IL-12, IFNγ, IL-21, IL-4, IL-5, IL-17A, IL-23, MIP-1α, fractalkine, and GM-CSF (**Fig. 3E**). This profile suggests that patients in cluster 2 exhibited a broader and more immunologically active systemic cytokine milieu, involving simultaneous activation of inflammatory, Th1-, Th2-, and Th17-related pathways, as well as enhanced chemokine and growth factor signaling. In contrast, neoadjuvant therapy and higher nodal status, which are characteristic of patients in cluster 1, may be associated with a dampened systemic immune response, as reflected by reduced cytokine levels. We next performed hierarchical correlation analysis to explore the relationships between soluble mediators and immune cell populations. This analysis showed that the cytokines elevated in cluster 2 positively correlated with B-cell subsets (**Fig. 3F**), which is consistent with the increased abundance of total B cells and multiple B-cell subpopulations previously observed in this cluster. Another prominent pattern of positive correlation was observed for CD4+ T-cell populations, which were associated with a broad range of the measured soluble factors. When cytokine levels were analyzed according to breast cancer subtype, only limited differences were detected. Specifically, significant differences were observed for GM-CSF and IFNγ, both of which showed the lowest levels in TNBC compared with luminal subtypes (**Fig. 3G**, **Fig. S8**). Consistent with this observation, correlation analysis across all patients revealed a strong positive relationship between GM-CSF and IFNγ, irrespective of cluster assignment.

### Mass cytometry reveals heterogeneity of tumor architecture in breast cancer

Breast tumors are composed not only of malignant epithelial cells, but also of stromal and immune cell populations that together shape the tumor microenvironment (9, 39). The relative abundance of these cellular compartments may influence tumor behavior, disease progression, and response to treatment. We therefore used mass cytometry to characterize the cellular architecture of 55 breast cancer tissues at the single-cell level and to assess its variability across molecular subtypes and clinicopathological features. Tumor samples were first dissociated into single-cell suspensions and stained with an antibody panel designed to identify fibroblasts, epithelial cells, endothelial cells, and immune cells (**Table S1**). Only samples yielding more than 10,000 cells were included in the analysis (**Fig. S9A**). Stratification of patients according to molecular subtype, including luminal A, luminal B HER2-, luminal B HER2+, HER2+ non-luminal, and triple-negative breast cancer, did not reveal any obvious trends or consistent patterns in the relative abundance of the major cellular compartments (**Fig. 4A, Fig. S9B).** Apoptotic cells accounted for approximately 5% to 25% of all cells. Among non-apoptotic cells, the largest populations were epithelial cells and other non-immune cells that could not be further classified, likely including adipocytes. Immune cells and fibroblasts also represented substantial fractions of the tumor tissue, whereas endothelial cells constituted the least abundant population. Overall, tumor architecture was highly heterogeneous and did not appear to be determined by molecular subtype. Similarly, no clear subtype-related pattern was observed when the composition of tumor-infiltrating immune cells, including granulocytes, B cells, T cells, NK cells, and macrophages, was examined at first glance (**Fig. 4A**). Unsupervised clustering, however, identified six patient clusters based on overall tumor architecture and five clusters based on immune cell infiltration patterns (**Fig. S9C,D**). For each sample, mass cytometry also enabled reconstruction of the tumor cellular landscape in the form of viSNE maps (**Fig. S10, S11**). These maps demonstrated substantial interpatient heterogeneity, even among tumors belonging to the same molecular subtype. For example, two patients with luminal B HER2-breast cancer (#12 and #55) displayed markedly different tissue organization: sample #12 was dominated by epithelial cells and fibroblasts with almost no immune infiltration, whereas sample #55 showed extensive immune infiltration, particularly by CD3+ T cells and B cells (**Fig. 4B**). Quantitative comparison of the relative abundance of major tumor cell populations across molecular subtypes did not reveal statistically significant differences (**Fig. 4C**). Notably, however, ER-positive tumors contained a significantly higher proportion of epithelial cells and a lower proportion of endothelial cells. In contrast, the abundance of immune cells and fibroblasts did not differ according to ER status (**Fig. 4D**). No significant differences in the abundance of major tumor-associated cell populations were observed in relation to HER2 status, Ki67 level, tumor stage, or neoadjuvant treatment (**Fig. S12**), although tumors with higher Ki67 showed a tendency toward increased immune infiltration. Further analysis focused on immune cells within the tumor microenvironment likewise showed no significant differences in the abundance of individual immune cell subsets across molecular subtypes (**Fig. 4E**). In contrast, stratification by neoadjuvant treatment revealed clear differences: patients who had received neoadjuvant therapy had a higher proportion of granulocytes and a lower proportion of T cells than untreated patients (**Fig. 4F**). Moreover, the previously noted association between higher Ki67 levels and increased immune infiltration appeared to be primarily driven by granulocytes (**Fig. S13**). No additional significant associations between immune cell abundance and HER2 status, ER status, or tumor stage were observed. Together, these mass cytometry data indicate that, regardless of clinical subtype, breast tumors are composed predominantly of epithelial cells. Immune cells also represent a substantial component of the tumor microenvironment, although their abundance varies considerably between patients. Overall, these findings are consistent with previous reports describing the pronounced heterogeneity of breast tumor architecture (9, 13).

**Figure 4.**
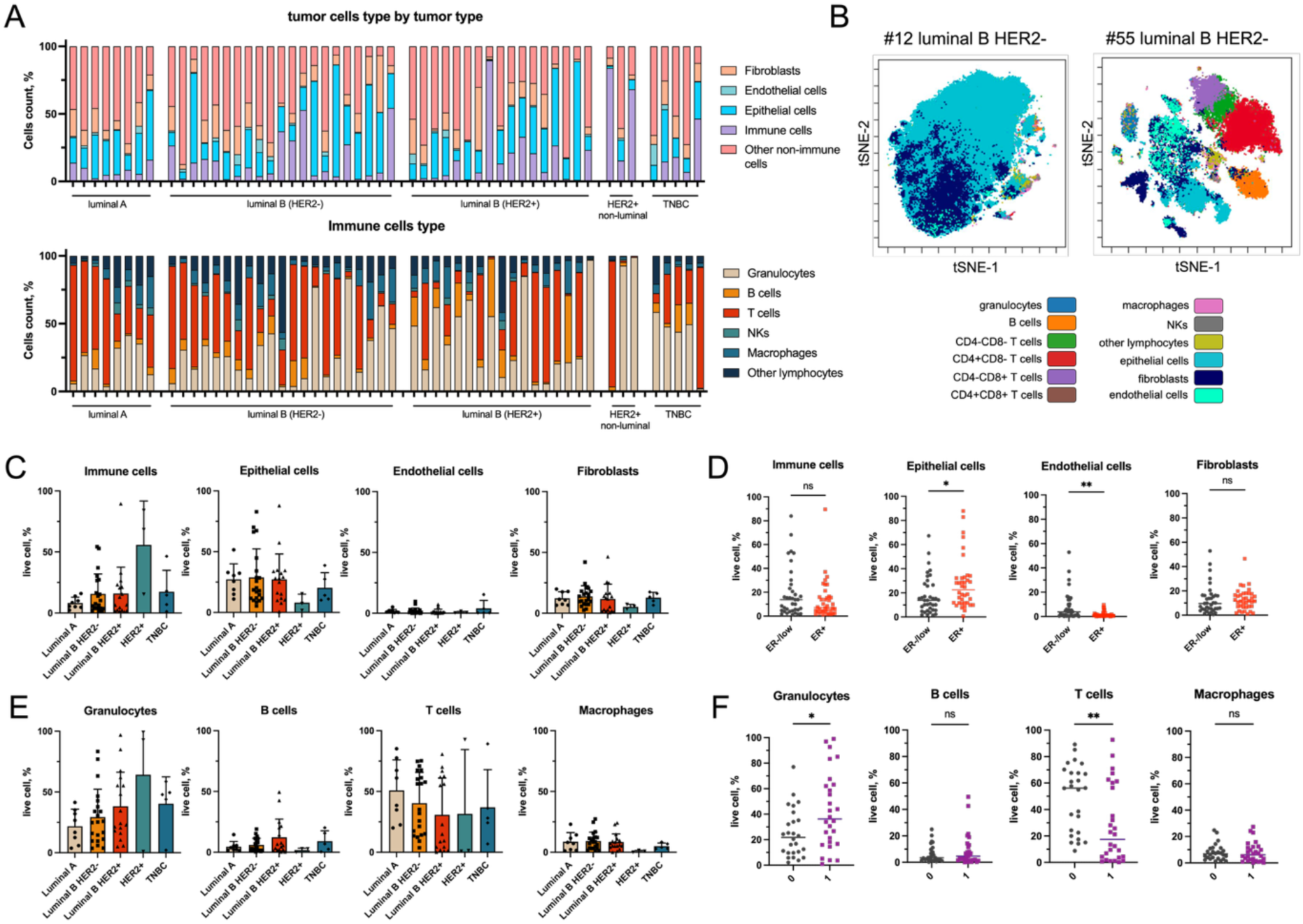
Analysis of the tumor cell landscape in breast cancer. **A** Quantitative analysis of major cell populations, including fibroblasts, endothelial cells, epithelial cells, immune cells, and other non-immune cells (top), as well as immune cell subtypes, including granulocytes, B cells, T cells, NK cells, macrophages, and other lymphocytes (bottom), presented as percentages for all patients stratified by breast cancer subtype. The data demonstrate substantial variability in tumor cell composition across patients and molecular subtypes. **B** Representative viSNE plots of two luminal B HER2-patients (#12 and #55), illustrating a tumor sample enriched in epithelial cells (#12) and a tumor sample enriched in infiltrating immune cells (#55). **C, D** Differences in the proportions of immune cells, epithelial cells, endothelial cells, and fibroblasts between patients stratified by molecular subtype **C** or ER status **D**. **E, F** Differences in the proportions of granulocytes, B cells, T cells, and macrophages between patients stratified by molecular subtype **E** or neoadjuvant treatment **F**. Statistical significance for all analyses was assessed using Welch’s t-test. * P < 0.05, ** P < 0.005, *** P < 0.0005.

### Protease and cystatin expression across tumor cell populations in breast cancer

In breast cancer, lysosomal cathepsins and legumain are expressed not only by malignant epithelial cells, but also by tumor-associated immune and stromal populations, indicating that their biological roles are strongly dependent on cellular context (40, 41). We therefore analyzed the expression of cathepsins B, L, S, and D, legumain, and their endogenous inhibitors cystatins B, C, and E/M at single-cell resolution in order to define how proteolytic programs are distributed across distinct cellular compartments of the tumor microenvironment (**Fig. 5A**). Single-cell analysis of cathepsin B, one of the best-characterized lysosomal cathepsins in cancer, showed that its expression was most prominent in CD4+ and CD8+ T cells, and was also clearly detectable in B cells and macrophages (**Fig. 5B**). Thus, cathepsin B was enriched primarily in immune cells rather than in the non-immune compartments of the tumor tissue. Interestingly, cathepsin L showed a similar pattern, as did cystatin C, a major endogenous inhibitor of cathepsins B and L (**Fig. S14A**). In contrast, cathepsins S and D displayed a related but distinct expression profile, with relatively broad distribution across most immune cell populations, although with lower representation in granulocytes. A different pattern was observed for legumain and its endogenous inhibitor cystatin E/M, which showed strong co-localization across cell populations, suggesting a particularly close biological relationship between these two proteins. Hierarchical clustering confirmed these observations, grouping legumain with cystatin E/M, cathepsins D and S into a second cluster, and cathepsins B and L together with cystatins C and B into a third cluster (**Fig. 5C**). Because cathepsin expression was particularly prominent in CD8+ T cells, we next examined the relationships among individual proteases and cystatins within this compartment. This is biologically relevant because lysosomal proteases in CD8+ T cells may reflect their activation state, intracellular turnover, and metabolic adaptation within the tumor microenvironment rather than direct extracellular proteolysis (42). In addition, CD8+ T cells are key mediators of anti-tumor immunity, and protease-related programs in these cells may therefore provide insight into their functional state within the tumor (43). Correlation analysis revealed the strongest associations between cathepsins and cystatins, as well as between legumain and cystatin E/M, indicating the presence of a robust protease-inhibitor axis (**Fig. 5D).** Among the enzymes themselves, cathepsins L and B, which belonged to the same hierarchical cluster, showed a moderate positive correlation across patients (ρ = 0.45), whereas cathepsins S and D showed little or no correlation with each other (**Fig. 5E**). Notably, expression of these enzymes in CD8+ T cells appeared to be largely independent of breast cancer subtype and prior neoadjuvant treatment, suggesting that this proteolytic program may reflect a relatively stable immune-cell intrinsic state rather than a feature tightly linked to conventional clinical classification. Consistent with these observations, viSNE analysis of tumors with high immune-cell content showed that cathepsins B and L were strongly expressed in both CD3+CD4+ and CD3+CD8+ T cells and displayed marked co-occurrence, whereas cathepsins S and D were less strongly associated with these populations (**Fig. 5F,G**). Another immune population of particular interest in the tumor microenvironment is macrophages, which are widely recognized as major regulators of tumor progression, matrix remodeling, angiogenesis, and immune suppression (44). Tumor-associated macrophages have also been closely linked to lysosomal activity and protease-dependent remodeling processes, making them a biologically relevant compartment for the analysis of cathepsins and their inhibitors (45). In macrophages, the correlation matrix again demonstrated a strong relationship between cathepsins L and B and cystatin C, as well as between legumain and cystatin E/M (**Fig. 5H**), indicating that these protease-inhibitor relationships are not restricted to lymphoid cells but extend to myeloid populations within the tumor microenvironment. The proportion of macrophages within tumor tissue, expressed as a fraction of all tumor-derived cells, ranged from 0% to 3%. The overall macrophage content did not differ significantly among molecular breast cancer subtypes (**Fig. 5I**, **S14B**). However, macrophage abundance did vary according to HER2 status, as patients with the highest HER2 expression by IHC (3+) had a significantly lower proportion of macrophages than the remaining patients. Similarly, patients with advanced-stage tumors showed lower macrophage abundance than those with early-stage disease (**Fig. S14B**). No additional patient stratification yielded significant macrophage-related differences. Interestingly, cathepsin B expression in macrophages was also lower in patients with advanced-stage tumors than in those with early-stage disease (**Fig. 5I**). This effect was not observed for the remaining proteins. Although the median expression of the other enzymes in macrophages also tended to be lower in late-stage disease, these differences did not reach statistical significance, which may reflect either a weaker biological effect or limited statistical power. As expected, high cathepsin B expression in macrophages strongly correlated with cystatin C expression (**Fig. 5J**), regardless of tumor subtype. Similarly strong relationships were observed for cathepsin L and cystatin C, as well as for legumain and cystatin E/M. Taken together, these results show that protease expression in breast tumors is organized into reproducible, cell type-associated programs rather than being randomly distributed across the tumor microenvironment. Across both lymphoid and myeloid compartments, the strongest and most consistent relationships were observed between cathepsins B/L and cystatin C, and between legumain and cystatin E/M, indicating the presence of conserved protease–inhibitor axes. At the same time, the clinical associations were relatively limited, suggesting that the distribution of these proteolytic enzymes is driven more strongly by cellular identity and functional state than by conventional clinicopathological classification alone.

**Figure 5.**
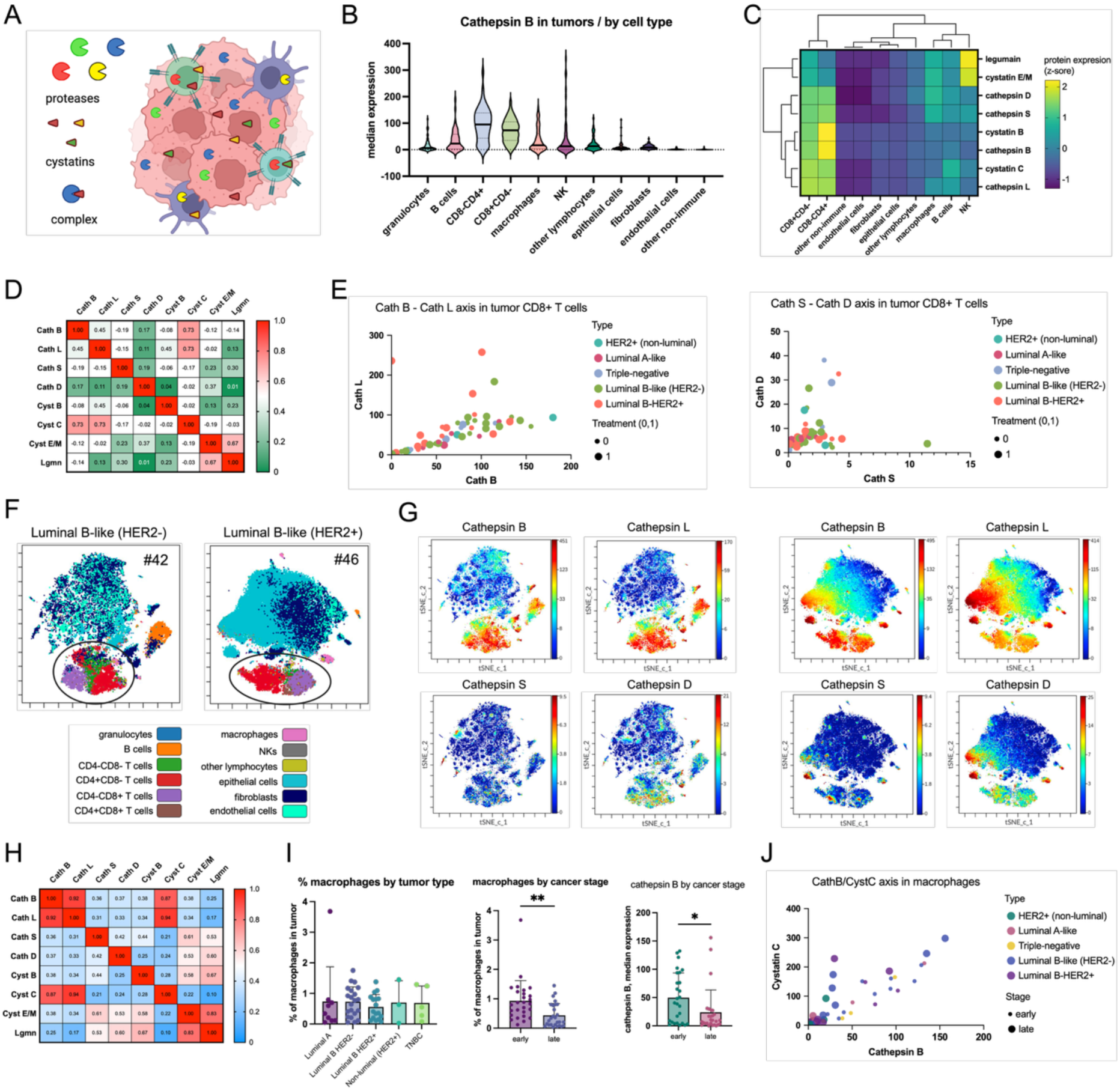
Analysis of protease expression in breast cancer samples. **A** Schematic representation of proteases and their endogenous inhibitors in cancer cells and cancer-associated cell populations. **B** Violin plots showing cathepsin B expression across individual cell types identified within tumor samples from all patients. **C** Hierarchical clustering of lysosomal proteases (cathepsins B, L, S, D, and legumain) and their endogenous inhibitors (cystatins B, C, and E/M) across the different cell types identified within tumor samples. **D** Correlation matrix of protease and inhibitor expression in CD8+ T cells. **E** Correlation plots showing the relationships between cathepsin B and cathepsin L (ρ = 0.45) and between cathepsin S and cathepsin D (ρ = 0.17) in CD8+ T cells, annotated by breast cancer subtype and neoadjuvant treatment status (0, 1). **F** Representative viSNE plots of two luminal B-like (HER2-) tumor samples (#42 and #46) enriched in CD3+ T cells. **G** viSNE analysis of samples #42 (left) and #46 (right) from patients with luminal B-like (HER2−) breast cancer, showing high expression of cathepsins B and L, moderate expression of cathepsin D, and low expression of cathepsin S in CD3+ T cells within tumor samples. **H** Correlation matrix of protease and inhibitor expression in macrophages. **I** Differences in macrophage abundance (percentage of tumor cells) in tumors from patients stratified by breast cancer subtype or tumor stage. Differences in cathepsin B expression in macrophages stratified by breast cancer subtype are also shown. **J** Correlation between cathepsin B and cystatin C expression in macrophages (ρ = 0.92), annotated by breast cancer subtype (colors) and tumor stage (bubble size). Statistical significance for all analyses was assessed using Welch’s t-test. * P < 0.05, ** P < 0.005, *** P < 0.0005.

### Analysis of protease expression in epithelial cells in the context of ADC development

Antibody-drug conjugates (ADCs) are an emerging class of anticancer therapeutics that combine the selectivity of monoclonal antibodies with the cytotoxic activity of highly potent payloads (46). In breast cancer, ADC-based treatment has become particularly important in HER2-positive disease, with trastuzumab emtansine and trastuzumab deruxtecan serving as representative examples (47, 48), while TROP-2-targeting ADCs have further expanded the clinical relevance of this strategy (49). Classically, ADCs act through antigen binding at the tumor cell surface, followed by internalization, trafficking to endolysosomal compartments, and intracellular linker cleavage, which releases the active payload (**Figure 6A**) (50). Increasing attention, however, has been directed toward protease-sensitive linker systems, including the widely used Val-Cit motif, which is hydrolyzed by multiple lysosomal cysteine cathepsins (51), as well as next-generation linkers designed for greater cathepsin selectivity (52, 53). Because HER2-directed ADCs primarily act within epithelial tumor cells that express the target antigen, we next focused on the epithelial compartment of breast tumors and examined the expression of cathepsins, legumain, and cystatins in individual patients. Hierarchical clustering identified four patient groups with distinct proteolytic profiles (**Fig. 6B**). The first cluster was characterized by globally low expression of cathepsins, legumain, and cystatins. The second showed moderate expression of legumain, cathepsin S, and cystatin E/M. The third was defined by high expression of cathepsins L and B together with cystatin C, whereas the fourth displayed either very high expression of cathepsins L and B or a profile dominated by high legumain and cathepsin D. Importantly, high protease expression did not consistently coincide with high HER2 expression by IHC, which is highly relevant for the design of HER2-targeting ADCs that depend on lysosomal proteolysis for efficient payload release (**Fig. 6B**). Because the abundance of epithelial cells within tumor tissue may influence the effective targetable compartment for ADC therapy, we next evaluated epithelial cell content across the cohort. The percentage of epithelial cells did not differ significantly with respect to molecular subtype, ER status, PR status, HER2 status, tumor stage, neoadjuvant treatment, or tumor grade (**Fig. S15A**). These findings suggest that tumors with different clinical and architectural characteristics may still be potentially susceptible to ADC-based treatment, provided that the epithelial compartment expresses both the target antigen and a compatible proteolytic machinery. Further analysis of cathepsins B and L, as the principal candidate enzymes for intracellular ADC activation, showed that both strongly correlated with cystatin C and with each other (**Fig. 6C**). In parallel, legumain again showed a strong association with cystatin E/M in epithelial cells, consistent with the protease-inhibitor relationships observed in other tumor cell populations (**Fig. S15B**). Because cathepsins B and L did not show a linear relationship with HER2 expression, we considered that a clinically relevant stratification strategy should incorporate both target expression and protease abundance. We therefore developed a scoring approach to identify patients whose epithelial tumor cells simultaneously displayed high HER2 expression and high levels of cathepsin B or cathepsin L. For viSNE analysis, we selected five patients with high HER2 expression by IHC (scores 2-3), including cases with high cathepsin B/L expression (#28, #34, and #46) and low cathepsin B/L expression (#47 and #65) (**Fig. 6D**). Single-cell viSNE maps confirmed strong co-localization of cathepsins B and L while also demonstrating that high HER2 expression did not necessarily predict high protease expression. Notably, cathepsin B expression in epithelial cells varied markedly across the cohort, spanning approximately two orders of magnitude (**Fig. 6E**). The median expression value of cathepsin B was 5.36, allowing classification of patients into cathepsin B-high and cathepsin B-low groups. When this stratification was combined with the clinical HER2 score, dichotomized as HER2-low (0–1) or HER2-high (2–3), 7 patients (13%) were identified as cathepsin B-high/HER2-high in epithelial cells, whereas 16 patients (29%) belonged to the cathepsin B-low/HER2-low group. These data indicate that the subgroup of patients who are mechanistically optimal candidates for HER2-targeting, cathepsin B-cleavable ADCs may be substantially narrower than HER2 status alone would suggest. A similar analysis performed for cathepsin L yielded comparable results. Using the cohort median of 7.58 as the cut-off, 6 patients were classified as cathepsin L-high/HER2-high (**Fig. 6F**). Because many clinically used protease-cleavable ADCs rely on linkers such as Val-Cit, which can be hydrolyzed by multiple lysosomal hydrolases rather than a single protease (54), we also examined LAMP-1 expression. LAMP-1 is a lysosomal membrane protein widely used as a marker of lysosomal abundance and endolysosomal activity (55), and thus provides an indirect readout of the intracellular compartment in which many ADCs undergo processing. In this context, LAMP-1 may be informative not only as a marker of lysosomal capacity, but also as a surrogate indicator of the cellular environment permissive for linker cleavage and payload release. LAMP-1 correlated well with cathepsin expression, and this analysis identified 10 patients as LAMP-1-high/HER2-high, suggesting that a LAMP-1/HER2 scoring strategy may represent a useful alternative to cathepsin B- or cathepsin L-based stratification (**Fig. 6F**). We next asked whether these observations could be recapitulated in experimental breast cancer models. Using the same antibody panel, we profiled three breast cancer cell lines representing distinct molecular subtypes: MDA-MB-231 (triple-negative), MCF-7 (luminal A, ER+/PR+), and BT-474 (HER2-positive, luminal B-like). The results confirmed substantial cathepsin expression across all three models together with the expected cell line-specific expression of ER, PR, and HER2 (**Fig. 6G**). We then evaluated cell death induced by incubation with ADCs of the general format anti-HER2 (trastuzumab)-peptide-PABC-MMAE, in which MMAE served as the antimitotic payload (**Fig. 6H**). Three conjugates were tested: a pan-cathepsin-cleavable ADC containing the classical Val-Cit linker, a cathepsin B-selective ADC containing the Cha-Leu-hSer(Bzl)-Gln sequence, and a cathepsin L-selective ADC containing the His-dThr-Phe(F_5_)-Cys(Bzl) sequence (52). BT-474 cells, which express high levels of HER2, responded strongly to all ADC constructs. In contrast, MDA-MB-231 and MCF-7 cells, which lack HER2 overexpression, were largely insensitive to HER2-targeting ADCs. A modest degree of toxicity was observed for the cathepsin B-selective ADC, likely reflecting the fact that cathepsin B can also be present extracellularly (56, 57), thereby enabling some level of extracellular linker cleavage and premature payload release displayin bystandar effect. Taken together, these findings support the concept that effective ADC action depends not only on the presence of a surface target such as HER2, but also on the intracellular proteolytic context of epithelial tumor cells. Our data suggest that integrating target expression with protease or lysosomal scoring may provide a more mechanistically informed framework for patient stratification than HER2 status alone. More broadly, these results support the development of next-generation ADC selection strategies based on biomarker-protease pairing, which may help identify the subgroup of patients most likely to benefit from protease-activated targeted therapy.

**Figure 6.**
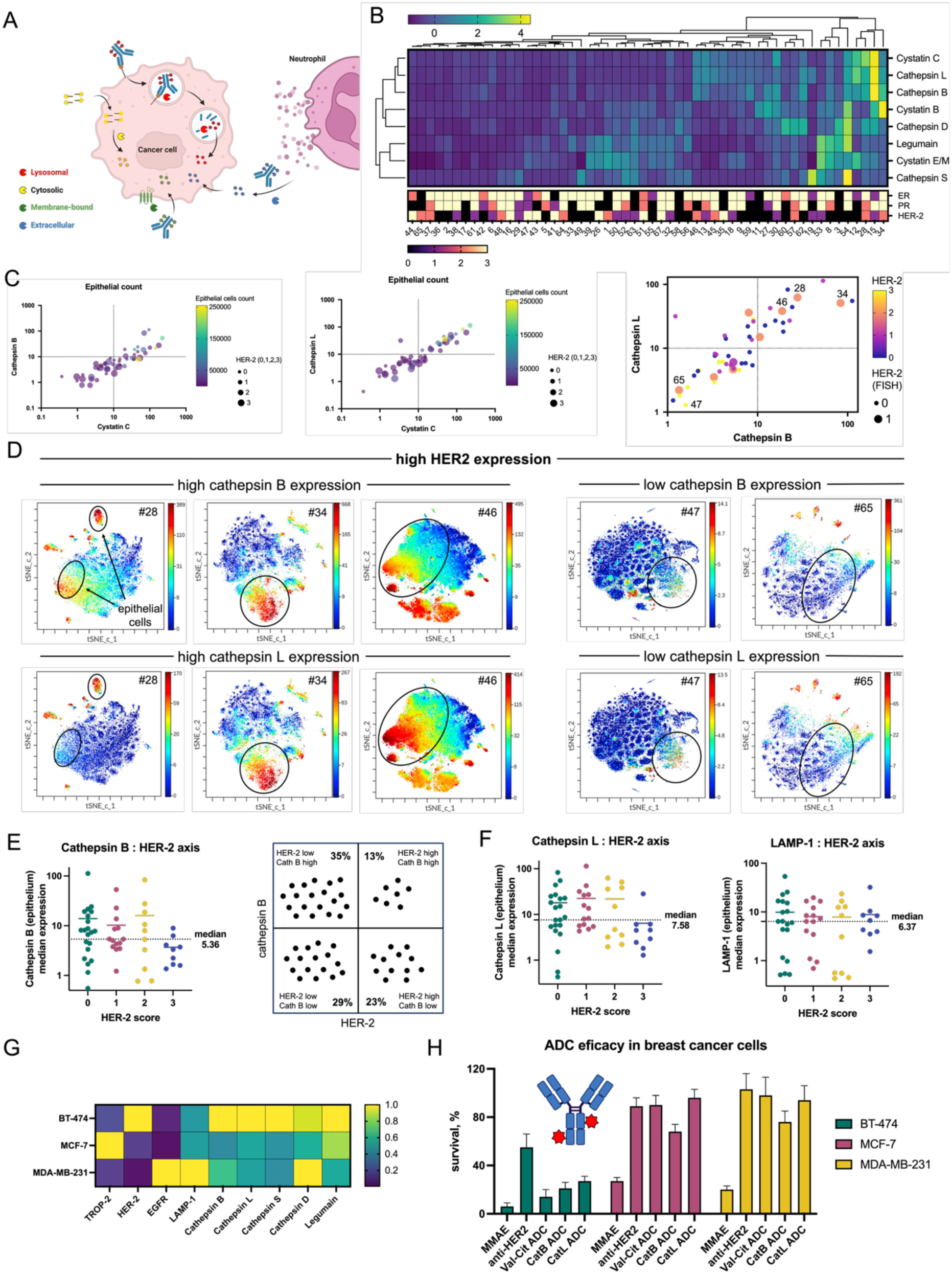
Identification of the optimal biomarker-protease pair for the development of antibody-drug conjugates. **A** Schematic overview of ADC mechanisms of action. The classical pathway of ADC activity involves binding to tumor cell surface antigens, followed by internalization and lysosomal hydrolysis, leading to release of the active payload. Alternative mechanisms include extracellular activation by proteases derived from tumor or tumor-infiltrating cells, as well as cytosolic activation by intracellular proteases. **B** Hierarchical clustering of patients based on the expression of proteases and their inhibitors in epithelial cells within tumor samples. ER, PR, and HER2 status are indicated below the heatmap for each patient. **C** Correlation plots showing the relationships between cystatin C and cathepsin B, and between cystatin C and cathepsin L, in epithelial cells, annotated by epithelial cell abundance (color) and HER2 status (bubble size). The correlation between cathepsin B and cathepsin L expression in epithelial cells is also shown, annotated by HER2 IHC status (color) and HER2 FISH status (bubble size). **D** viSNE plots of tumor samples from selected patients (#28, #34, #46, #47, and #65) with high HER2 expression by IHC, illustrating cases with high cathepsin B/L expression in epithelial cells (left) or low expression of these proteases (right). Epithelial cells are indicated by black circles on the viSNE plots. **E** Dot plot showing cathepsin B expression in epithelial cells from patients stratified by HER2 IHC status (0, 1, 2, 3). The median value (5.36) is indicated by a dotted line, defining cathepsin B-high and cathepsin B-low groups. These data were used to generate a cathepsin B/HER2 scoring matrix, which identified 7 patients (13%) with high cathepsin B and HER2 expression in epithelial cells and 16 patients (29%) with low cathepsin B and HER2 expression. **F** Dot plots showing cathepsin L (left) and LAMP-1 (right) expression in epithelial cells from patients stratified by HER2 IHC status (0, 1, 2, 3). Median values of 7.58 for cathepsin L and 6.37 for LAMP-1 are indicated by dotted lines. **G** Expression of cathepsins, legumain, and selected breast cancer markers (HER2, TROP-2, EGFR, and LAMP-1) in three breast cancer cell lines representing distinct disease subtypes: MDA-MB-231 (triple-negative), MCF-7 (luminal A, ER+/PR+), and BT-474 (HER2+, luminal B-like). Expression is shown as scaled z-scores from 0 to 1. **H** Efficacy of anti-HER2 antibody-drug conjugates (trastuzumab-peptide-PABC-MMAE) across three breast cancer cell lines. MMAE denotes free monomethyl auristatin E; anti-HER2 denotes trastuzumab; Val-Cit represents a classical ADC containing the pan-cathepsin-cleavable dipeptide linker; CatB ADC contains a cathepsin B-selective Cha-Leu-hSer(Bzl)-Gln tetrapeptide linker; and CatL ADC contains a cathepsin L-selective His-dThr-Phe(F5)-Cys(Bzl) tetrapeptide linker.

### Spatial analysis of proteases in breast cancer by imaging mass cytometry

Although single-cell CyTOF analysis provided detailed information on protease expression across individual tumor and immune cell populations, it did not preserve the spatial context needed to determine how these proteins are distributed within intact tissue or how they relate to epithelial and immune compartments in situ. We therefore used imaging mass cytometry to define the spatial localization of proteases in breast cancer tissues and to assess their association with epithelial, stromal, and immune cell populations. Selected patient samples were processed into formalin-fixed, paraffin-embedded tissue sections and analyzed by imaging mass cytometry to resolve the spatial organization of breast tumors at the tumor margin, where malignant cells interact most directly with the surrounding microenvironment (**Fig. 7A**). To define tissue architecture and cellular composition, we included structural markers of stromal and epithelial organization, including α-SMA, pan-keratin, vimentin, and collagen I, together with immune markers identifying CD4+ and CD8+ T cells, CD20+ B cells, and CD68+ macrophages. This marker panel was selected to capture the major cellular and extracellular compartments known to shape tumor progression, immune response, and local tissue remodeling in breast cancer. The tumor region of sample MASP110, invasive breast carcinoma of no special type (NST), showed moderate desmoplastic stroma, characterized by strong vimentin and collagen I signals and only low expression of α-SMA (**Fig. 7B, left).** Vimentin expression was particularly prominent around adipose tissue and at the tumor margins, whereas pan-keratin, marking epithelial cells, defined a distinct tumor cell compartment. The same section also showed widespread immune cell infiltration throughout the tissue (**Fig. 7B, middle**). CD20+ B cells formed dense and well-defined aggregates, particularly at tumor margins adjacent to adipose tissue, but were also present within the tumor core. CD68+ macrophages were enriched both at the tumor border and in internal tumor regions. CD4+ and CD8+ T cells also showed partial co-localization, although their spatial distribution remained distinct from that of CD68+ macrophages and CD20+ B cells. Analysis of protease localization revealed that cathepsin D was highly expressed throughout the section, forming the dominant protease signal (**Fig. 7B, right**). In contrast, cathepsin B showed a more restricted pattern, with enrichment around adipocyte-rich regions, whereas cathepsin S was sparse and largely confined to the tumor margins and a few internal foci. Spatial comparison of protease and immune markers suggested that cathepsin B was more closely associated with CD68+ macrophages, while cathepsin D showed stronger co-localization with CD20+ B cells. An additional sample of invasive breast carcinoma of no special type (NST) showed minimal α-SMA expression and a clear spatial separation between pan-keratin-rich epithelial regions and collagen I/vimentin-rich stromal regions (**Fig. S16, left**). This section also displayed pronounced immune infiltration, with CD4+ T cells and CD20+ B cells predominating over CD8+ T cells and CD68+ macrophages. As in the previous cases, cathepsin D was the most abundant protease throughout the section, whereas cathepsin B was enriched in vimentin-rich and immune-infiltrated regions, particularly in association with CD68+ macrophages. Because cancer-associated adipocytes are increasingly recognized as active components of the breast tumor microenvironment that contribute to inflammation, extracellular matrix remodeling, and tumor progression (58), we also examined the corresponding cancer-associated adipocyte area (**Fig. S16, right**). This region showed extensive immune infiltration dominated by CD68+ macrophages and CD4+ T cells, together with strong collagen and vimentin signals and low α-SMA expression. In this area, both cathepsin D and cathepsin B were clearly detectable, with cathepsin B enriched in collagen-rich regions and showing partial co-localization with vimentin and CD8+ T cells. Next, we also analyzed a non-invasive breast carcinoma sample by IMC. A strong α-SMA signal was detected across most of the section, consistent with its role as a marker of the myoepithelial layer surrounding ductal structures, which is typically preserved in non-invasive disease (**Fig. 7C, left**). In this sample, α-SMA was interwoven with pan-keratin and collagen I, whereas vimentin expression was lower and remained largely confined to the tumor core, in contrast to the other analyzed cases. Overall, the tissue architecture appeared substantially more compact and organized than that observed in the invasive samples. The α-SMA-rich region showed only minimal immune cell infiltration (**Fig. 7C, middle**). CD68+ macrophages were located mainly at the tumor margin, while CD4+ and CD8+ T cells were present only in low numbers and CD20+ B cells were sparse. Analysis of protease distribution showed high cathepsin B accumulation along the tumor margin, where it formed dense aggregates (**Fig. 7C, right**). The α-SMA-rich region was also enriched in cathepsin S, whereas cathepsin D displayed a more diffuse distribution throughout the section. Together, these findings suggest that protease activity in non-invasive breast carcinoma is spatially compartmentalized, with distinct protease patterns associated with the tumor border and myoepithelial-rich regions. We also analyzed a rare case of mucinous breast carcinoma, a histological subtype typically associated with favorable clinical outcome and low metastatic potential (59). The section displayed the characteristic loose architecture of mucinous carcinoma, with epithelial nests appearing to float within abundant extracellular mucin and stromal regions enriched in collagen I and vimentin (**Fig. 7D**). Several distinct structural regions could be identified, including duct-like epithelial structures, irregular mucin-filled cavities lined by pan-keratin-positive cells, and lobular-like areas with prominent pan-keratin and α-SMA expression. Immune cells were distributed throughout the tissue, with focal accumulations of CD68+ macrophages, CD4+ and CD8+ T cells, and CD20+ B cells in selected regions. Cathepsin B showed the broadest and strongest distribution, with pronounced enrichment in immune-rich areas and along epithelial structures bordering mucin-filled lumina, whereas cathepsins D and S were present more diffusely and at lower intensity. Overall, these findings indicate that even in this rare histological subtype, protease localization remains spatially heterogeneous and is closely linked to both epithelial architecture and local immune infiltration. Finally, we asked whether IMC could also be used to localize active proteases using chemical probes. TOF-probes consist of three key elements: an electrophilic warhead that covalently binds the active site of the enzyme, a recognition sequence that confers protease selectivity, and a detectable metal isotope tag (**Fig. 7E**) (32). Although such probes have previously been used to detect protease activity in single cells by CyTOF and in slide-based cellular systems (32, 33), their application to IMC in human tissue samples has not been established. This approach is particularly relevant in the context of our study, because antibody-based detection reports total protein abundance but does not distinguish between inactive and catalytically active enzyme. That distinction is critical for protease-activated ADCs, where therapeutic efficacy depends not simply on protease expression, but on the presence of functionally active enzyme capable of linker cleavage. This issue is especially important in light of our single-cell data showing strong correlations between cathepsins and their endogenous inhibitors, including cathepsins B and L with cystatin C, and legumain with cystatin E/M. These relationships indicate that high protease expression cannot be assumed to reflect high proteolytic activity. We therefore reasoned that activity-based IMC could provide a more mechanistically informative way to assess whether a given tumor is permissive for protease-dependent ADC activation. In principle, such an approach could later be integrated with biomarker staining to generate tissue-level scoring systems that combine target expression with local protease activity. As a proof of concept, we used the cathepsin B-selective TOF-probe TOF-120, which showed high selectivity toward cathepsin B and no detectable inhibition of other cancer-associated proteases tested in our panel. This specificity is conferred by a selective tetrapeptide recognition sequence containing unnatural amino acids, Cha-Leu-hSer(Bzl)-Arg, linked to an AOMK warhead (**Fig. 7E**) (34). Breast cancer tissue sections stained with TOF-120 showed broad cathepsin B activity distributed throughout the tissue (**Fig. 7F**). As a control for nonspecific binding, an adjacent section of the same carcinoma was preincubated with the pan-cathepsin inhibitor E64 before TOF-120 staining. Under these conditions, probe-derived signal was nearly abolished, indicating that the observed labeling depended on active cathepsin binding rather than nonspecific tissue retention. To our knowledge, this represents the first demonstration of imaging proteolytic activity in human breast carcinoma by IMC. More broadly, this strategy may provide a valuable platform for simultaneous spatial analysis of protease activity and breast cancer biomarkers, with potential applications not only in ADC development but also in the study of protease function within the tumor microenvironment.

**Figure 7.**
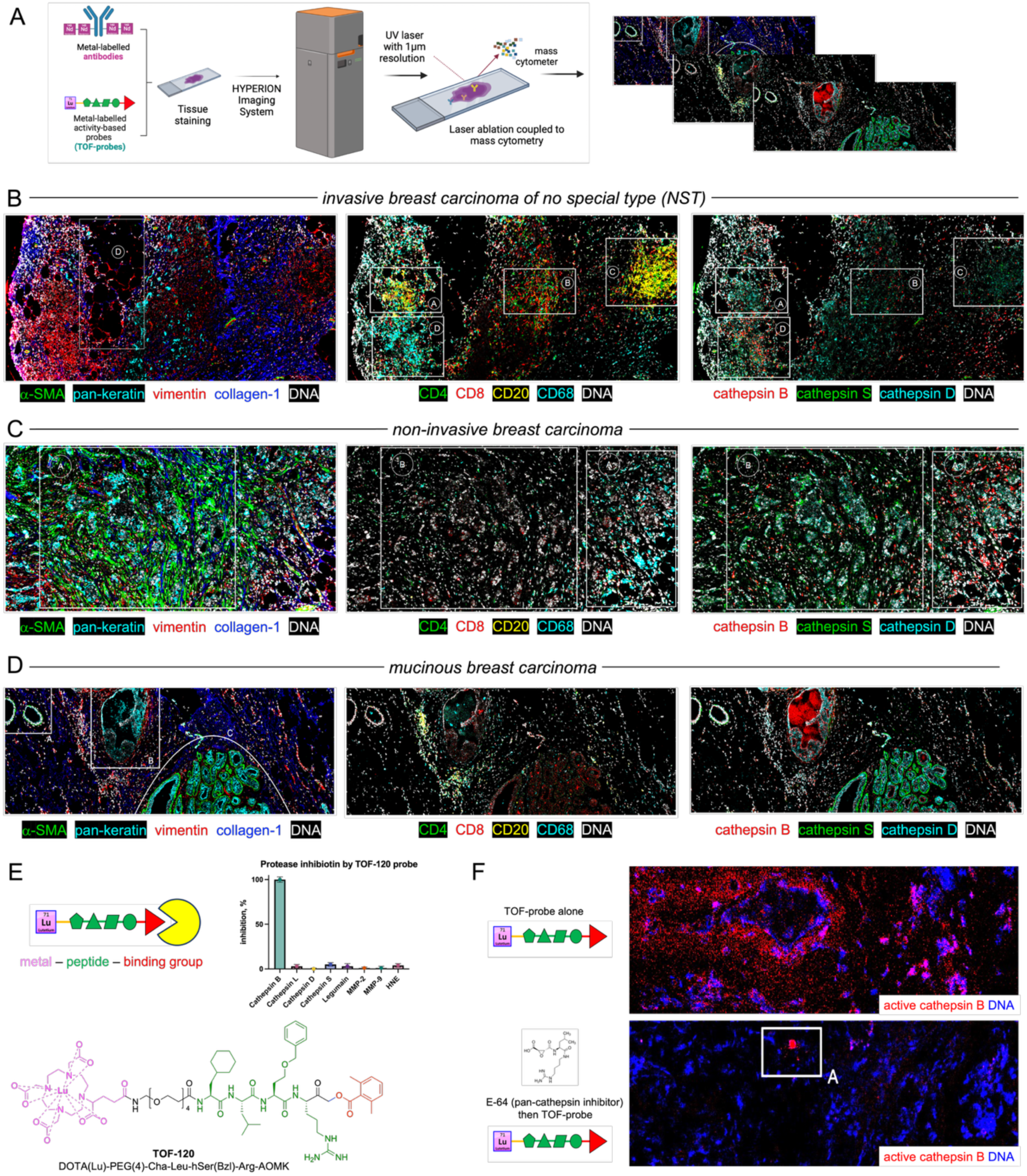
Protease localization in breast cancer by imaging mass cytometry. **A** General workflow for the detection of protease expression and activity by imaging mass cytometry (IMC). OCT-embedded tissues were labeled with an antibody cocktail containing markers of tissue structure, immune cells, and selected proteases. Selected sections were also stained with metal-tagged activity-based probes (TOF-probes) for the detection of active cathepsin B. **B-D** Architecture of representative samples of invasive breast carcinoma of no special type (NST) **B**, non-invasive breast carcinoma **C** and rare mucinous breast carcinoma **D**, visualized based on the spatial expression of structural markers (α-SMA, pan-keratin, vimentin, and collagen I) and infiltrating immune cells (CD4, CD8, CD20, and CD68). In addition, the samples were stained with antibodies against cathepsin B, cathepsin S, and cathepsin D. DNA is shown in white. Each marker was individually scaled to facilitate visualization. **E** General structure of a metal-tagged TOF-probe used for protease activity detection by mass cytometry. Selectivity of the cathepsin B TOF-probe (TOF-120) tested against a panel of cancer-associated proteases, including cathepsins B, L, D, and S, legumain, MMP-2, MMP-9, and HNE. The y-axis shows the percentage of protease inhibition. Chemical structure of the TOF-120 probe used for cathepsin B labeling: DOTA(Lu)-PEG(4)-Cha-Leu-hSer(Bzl)-Arg-AOMK. **F** Visualization of active cathepsin B in breast cancer tissue by IMC. The upper panel shows tissue stained with the TOF-probe alone, whereas the lower panel shows tissue preincubated with E64 (a pan-cathepsin inhibitor) followed by staining with TOF-120.

## Discussion

Our study takes a deliberately protease-centric view of breast cancer biology and places it in the context of the systemic immune state. Large single-cell and spatial atlases have already demonstrated that breast tumors form heterogeneous ecosystems in which malignant, immune, and stromal compartments vary strongly between patients and carry prognostic information (9). Rather than aiming to expand these atlases in breadth, we focused on two complementary questions: whether peripheral blood immune features and soluble mediators capture clinically relevant systemic states in breast cancer, and how lysosomal proteases and their endogenous inhibitors are distributed across tumor cell populations, at the single-cell and spatial levels, and what this implies for protease-activated therapeutic strategies. A key concept underlying the second part of the manuscript is that protease biology is inherently context dependent. Cathepsins operate within a dynamic “protease web” in which substrate access, activation, trafficking, extracellular acidification, and inhibition by cystatins collectively determine functional output (17). This is particularly relevant in tumors, where tumor-associated macrophages, fibroblasts, malignant epithelial cells, and extracellular matrix remodeling converge to create spatially restricted niches that can either promote or restrain tumor progression (60). Against this background, our results support three overarching messages: (1) peripheral immune composition and cytokine patterns identify distinct systemic states that co-segregate with treatment exposure and tumor aggressiveness; (2) cathepsins and cystatins form reproducible, cell-type-associated modules across the tumor ecosystem, with particularly informative patterns in macrophages/APCs and epithelial tumor cells; and (3) activity-resolved protease imaging is both feasible and mechanistically important, because expression alone cannot reliably report proteolytic competence in the presence of endogenous inhibition.

Peripheral blood represents a clinically accessible window into host-tumor interactions, capturing signals shaped by tumor burden, stromal remodeling, and therapy-induced immune modulation. In our cohort, immune-cell frequency patterns stratified patients into distinct clusters that strongly overlapped with neoadjuvant treatment exposure and nodal status, indicating that systemic immune features can capture clinically meaningful states even when tumor subtype alone provides limited separation (**Fig. 2**). This is consistent with the wider view that the tumor microenvironment and systemic immunity are dynamically coupled, and that stromal and immune compartments can be educated rather than simply recruited, contributing to disease progression and therapeutic response (39, 60). Importantly, the systemic cytokine milieu differed between clusters and suggested that one patient group maintained a broader, more immunologically active circulating profile, spanning inflammatory, Th1-, Th2-, and Th17-associated pathways together with chemokines and growth factor (**Fig. 3**). Such a pattern is biologically plausible in untreated settings where immune activation may reflect tumor-driven inflammation and ongoing immune surveillance. In contrast, neoadjuvant therapy is known to remodel both local and peripheral immunity, and can either enhance antitumor immune programs or dampen specific cytokine responses depending on regimen, timing, and tumor context (61). Although our dataset is cross-sectional, the strong association between systemic immune composition and treatment exposure underscores the need to treat therapy as a central covariate rather than a secondary annotation when interpreting blood-derived immune signals.

In peripheral immune cells, the distribution of cathepsins was dominated by cell identity, with the strongest signals in myeloid compartments (**Fig. 3**). This observation is consistent with broad mechanistic literature showing that cathepsins are integrated into endolysosomal functions in immune cells (including antigen processing, receptor recycling, and lysosome-mediated stress responses), and that changes in cathepsin activity are common in inflammation-associated states (37). The selective enrichment of cathepsin S in antigen-presenting lineages also fits well with its established role in MHC class II pathway maturation through invariant chain processing in dendritic cells and other APCs (62). A clinically relevant inference from our blood data is that protease expression signals and cytokine signals do not necessarily change in parallel. This decoupling is unsurprising given multilayer regulation: cathepsins are synthesized as proenzymes, require appropriate activation and trafficking, and are buffered by endogenous inhibitors such as cystatins (63). Therefore, blood-based protease abundance should be interpreted as a marker of proteolytic potential and cellular state, not as a direct surrogate of catalytic output. This interpretive constraint motivated our later activity-based analysis in tissue, and it supports a broader methodological recommendation - future blood-based stratification approaches that include proteases may benefit from integrating either activity readouts or orthogonal proxies of lysosomal state (e.g., LAMP-1-linked lysosomal capacity) rather than relying on abundance alone.

At the tumor single-cell level, we observed that cathepsin B was enriched in a subset of CD68⁺ TAMs and also detectable in a subset of epithelial tumor cells, whereas cathepsin D showed broad expression across many tumor cells and was also present in macrophages (**Fig. 5**, **Fig. 7**). This heterogeneous distribution aligns with the concept that cathepsins contribute to tumor progression through both tumor-intrinsic and microenvironmental (immune/stromal) mechanisms, including ECM turnover, invasive behavior, and therapy response modulation. Our data further suggest that cathepsin S is largely confined to APC-like populations and is often detectable at lower intensity than cathepsins B/D, consistent with its specialized immunological role. A central and recurring pattern in our single-cell analysis is the strong coupling between proteases and their endogenous inhibitors. Cathepsins B and L correlated with cystatin C, and legumain closely aligned with cystatin E/M, forming robust protease–inhibitor pairs across multiple cellular compartments. This is mechanistically important because cystatins can mask the functional meaning of high protease expression: high abundance may coexist with limited free activity, and conversely, modest expression can be functionally impactful if inhibition is relieved or trafficking changes. These observations support a model in which tumors do not simply upregulate protease expression, but rather tune proteolytic programs through coordinated regulation of enzymes and inhibitors - a feature that is likely shaped by both immune cell differentiation and the local biochemical environment (64). Notably, the lack of strong dependence of protease expression patterns on classical clinical stratifications in some compartments suggests that protease programs may reflect conserved cell-state modules, such as macrophage maturation/activation states, lysosomal biogenesis programs, rather than direct tumor subtype fingerprints. This resonates with microenvironment-focused frameworks in which stromal and immune education can create convergent pro-tumor phenotypes across genetically distinct tumors.

Imaging mass cytometry added a critical spatial dimension by showing that protease signals are not uniformly distributed across tissue but concentrate in architecturally and functionally distinct niches (**Fig. 7**). In invasive tumors, cathepsin D formed a dominant, broadly distributed signal, consistent with a pervasive lysosomal/proteolytic program in tumor cells, while cathepsin B was more restricted and frequently enriched in regions adjacent to adipocyte-rich areas and macrophage-infiltrated margins. Such patterns are consistent with the general notion that immune and stromal compartments shape, and are shaped by, localized remodeling at tumor margins, where invasion and tumor-stroma communication are most active (65). The observation that cathepsin B spatially aligns with CD68⁺ macrophages supports a TAM-linked proteolytic niche model, in which TAM-derived proteases act as microenvironmental effectors of invasion and tissue remodeling (66). However, antibody staining alone cannot resolve whether the detected enzymes are catalytically active, especially in light of the strong protease-cystatin coupling we observed. This limitation is well recognized in cathepsin biology. Expression-only measures can be difficult to interpret because they disregard inhibitor abundance, subcellular localization, and intratumoral heterogeneity of activity. To address this mechanistic gap, we implemented an activity-based imaging strategy using a mass cytometry-compatible TOF-probe (32). TOF-probes were previously introduced as lanthanide-tagged activity-based probes that enable multiplexed detection of protease activity by CyTOF and can be adapted to imaging mass cytometry, conceptually extending proteomics toward an “activome” readout (32). In our proof-of-concept tissue experiment, probe signal was effectively abrogated by pan-cathepsin blockade (E64), supporting specificity for active enzyme labeling and demonstrating feasibility of spatial activity mapping in human breast tumor sections. This advance aligns measurement with the biological quantity that determines protease-cleavable prodrug or linker processing, thereby strengthening mechanistic interpretation relative to expression-only data.

A major translational motivation for mapping epithelial protease programs is the growing clinical importance of antibody-drug conjugates and, specifically, the dependence of many ADCs on lysosomal processing for payload release. For example, T-DM1 relies on HER2-mediated internalization followed by lysosomal degradation of the antibody to release active DM1-containing metabolites (67). By contrast, multiple clinically used and investigational ADCs employ protease-cleavable peptide linkers, including the widely used Val-Cit-PABC motif and related designs (68, 69). More broadly, reviews have emphasized that the ADC field has been dominated by cathepsin-cleavable linker strategies and that activity-based probe approaches are increasingly relevant for diagnostic imaging and targeted delivery concepts (70). Two aspects of linker biology are particularly relevant for interpreting our data. First, while Val-Cit linkers were historically described as cathepsin B cleavable, accumulating evidence indicates that multiple lysosomal proteases can contribute to cleavage, including cathepsins S, L, and others, suggesting a degree of redundancy that may increase robustness but complicates attempts at strict single-enzyme selection (54, 71). Second, HER2 ADCs with alternative cleavable linkers, such as trastuzumab deruxtecan with a tetrapeptide linker, can be processed by lysosomal enzymes such as cathepsins B and L, underscoring the relevance of epithelial lysosomal protease context beyond HER2 expression alone. Our epithelial single-cell profiling directly supports a protease-informed stratification concept, in which high HER2 expression does not guarantee high cathepsin abundance (or activity), and conversely, high protease expression can occur in tumors with lower HER2 (**Fig. 5**). This decoupling implies that “target-only” selection is mechanistically incomplete for protease-activated ADCs. Instead, the optimal scenario is co-presence of sufficient surface target density for internalization and sufficient lysosomal proteolytic competence for efficient linker processing - ideally measured as activity rather than abundance given inhibitor effects. In this framework, our proposed scoring approaches (HER2 combined with cathepsin B/L or LAMP-1) represent a practical step toward functional patient selection that aligns biomarker readouts with ADC mechanism of action. Importantly, our *in vitro* experiments in breast cancer cell lines further support the expected dependence of HER2-directed ADC cytotoxicity on target expression, while also illustrating a key caveat of protease-cleavable designs. In settings where cathepsin activity is extracellular or pericellular, partial target-independent payload release may occur, potentially contributing to off-target toxicity or bystander effects.

## Conclusions

This study provides a cysteine cathepsins-centered, single-cell and spatial view of breast cancer that complements existing ecosystem atlases by connecting systemic immune states with intratumoral proteolytic programs. Across tumor compartments, cathepsin expression was strongly cell-type dependent and organized into reproducible protease–inhibitor modules, with cathepsin B enriched in subsets of CD68⁺ macrophages and some epithelial tumor cells, cathepsin D broadly distributed across tumor cells, and cathepsin S largely confined to APC-like populations. The strong coupling between cathepsins and cystatins highlights a key interpretive constraint, in which protease abundance alone cannot be assumed to reflect proteolytic activity. Building on mass cytometry-compatible activity-based probes, we demonstrate the feasibility of spatially resolving active protease signal in human breast tumor tissue, providing a mechanistically aligned readout for protease-dependent processes. From a translational perspective, our findings support the idea that protease-activated ADC efficacy depends on more than target expression: epithelial protease/lysosomal state and local protease activity can vary widely and may define a narrower subset of mechanistically optimal candidates for protease-cleavable HER2 ADCs. Combining target markers with protease abundance/activity or lysosomal surrogates (such as LAMP-1) offers a rational path toward protease-informed patient stratification and ADC design, and establishes a methodological foundation for future “activome-enabled” spatial tumor profiling.

## Limitation of the study

Several limitations should be considered when interpreting the present study. First, the cohort size and heterogeneity, especially in rarer subtypes, limit power for some subgroup analyses and prevent definitive linking of protease states to long-term clinical outcomes. Second, peripheral blood immune clustering is substantially confounded by treatment exposure; although this is biologically informative, it complicates attribution of systemic immune changes to tumor biology alone and motivates validation in treatment-naïve and longitudinal designs. Third, while we provide both expression-based and activity-based evidence, activity mapping was performed as a proof-of-concept for a single protease probe. Extending this to multiplex TOF-probe panels (cathepsin B/L, legumain, and potentially serine proteases) would enable more complete “activome” signatures that better match the redundancy of lysosomal cleavage mechanisms in ADC processing. Finally, because cathepsins can exert tumor-promoting or context-dependent functions, protease-informed therapy strategies will require careful evaluation of on-target efficacy against potential consequences of modulating proteolytic networks in immune and stromal compartments. Looking forward, the most impactful next steps are validation of protease-informed ADC scoring against treatment response (including pathological response to neoadjuvant therapy where available), integration of spatial activity mapping with multiplex tumor biomarker imaging to define “ADC-permissive niches,” and finally testing whether protease-rich TAM niches correlate with immune suppression states identified by broader ecosystem studies and whether they represent actionable combination targets.

## Data Availability

All data produced in the present study are available upon reasonable request to the authors

## Acknowledgements

This project was supported by the National Science Centre in Poland; grant SONATA UMO-2018/31/D/NZ5/02406 to MP and OPUS-LAP UMO-2020/39/I/NZ5/03104 to MP. We thank Prof. Boris Turk for providing recombinant cysteine cathepsins.

## Authors contribution

**N.Ć.-P.** designed the experiments, prepared antibody panels for mass cytometry, processed blood and breast cancer samples, designed and performed mass cytometry experiments, analyzed and interpreted the data. **N.M.-Ch.** designed and performed tissue visualization by IMC and analyze the IMC data. **O.G.** synthesized antibody-drug conjugates and performed ADC toxicity studies with breast cancer cell lines. **T.P.** and **J.M.** performed CyTOF data biostatistcial analysis. **J.N.** synthesized cathepsin B TOF-probes and performed kinetic analysis. **B.D.-K.** and **P.K.** collected and processed breast tumor samples. **M.S.** prepared the IMC slides and contributed to data analysis. **K.G.** designed the original metal-labeled antibody panel for tumor profiling and contributed to data analysis. **B.Sz.** and **R.M.** collected and processed breast tumor samples, supervised experimental design, and contributed to data interpretation. **M.P.** conceptualized and supervised the study, designed the experiments, provided funding and resources, and wrote the manuscript. All authors critically reviewed, revised, and approved the final manuscript.

