## Supplemental file for "Single-cell and spatial profiling of cysteine cathepsins identifies tumor states relevant to antibody-drug conjugates in breast cancer"

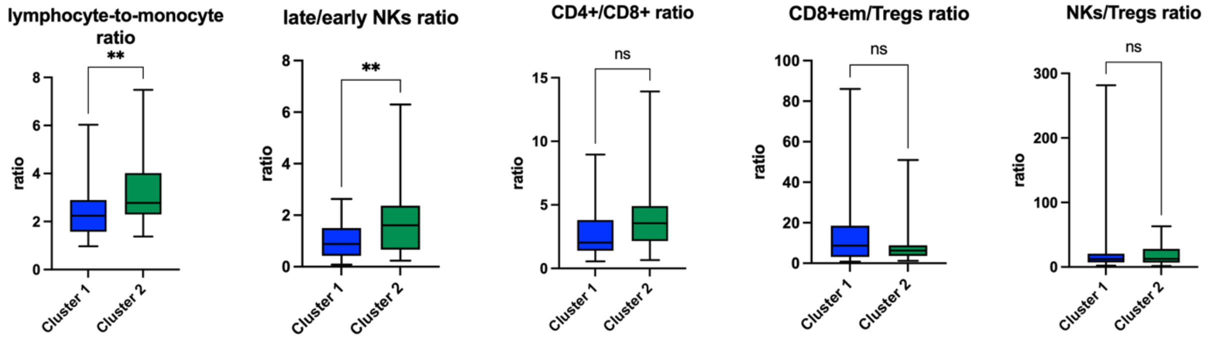


**Figure S1 Differences in selected immune cell ratios** between patients assigned to cluster 1 and cluster 2. The lymphocyte-to-monocyte ratio and the late/early NK cell ratio showed statistically significant differences, whereas no significant differences were observed for the CD4+/CD8+ ratio, CD8+ effector/Treg ratio, and NK/Treg ratio. Statistical significance was assessed using Welch's t-test. * $P < 0.05$, ** $P < 0.005$, *** $P < 0.0005$

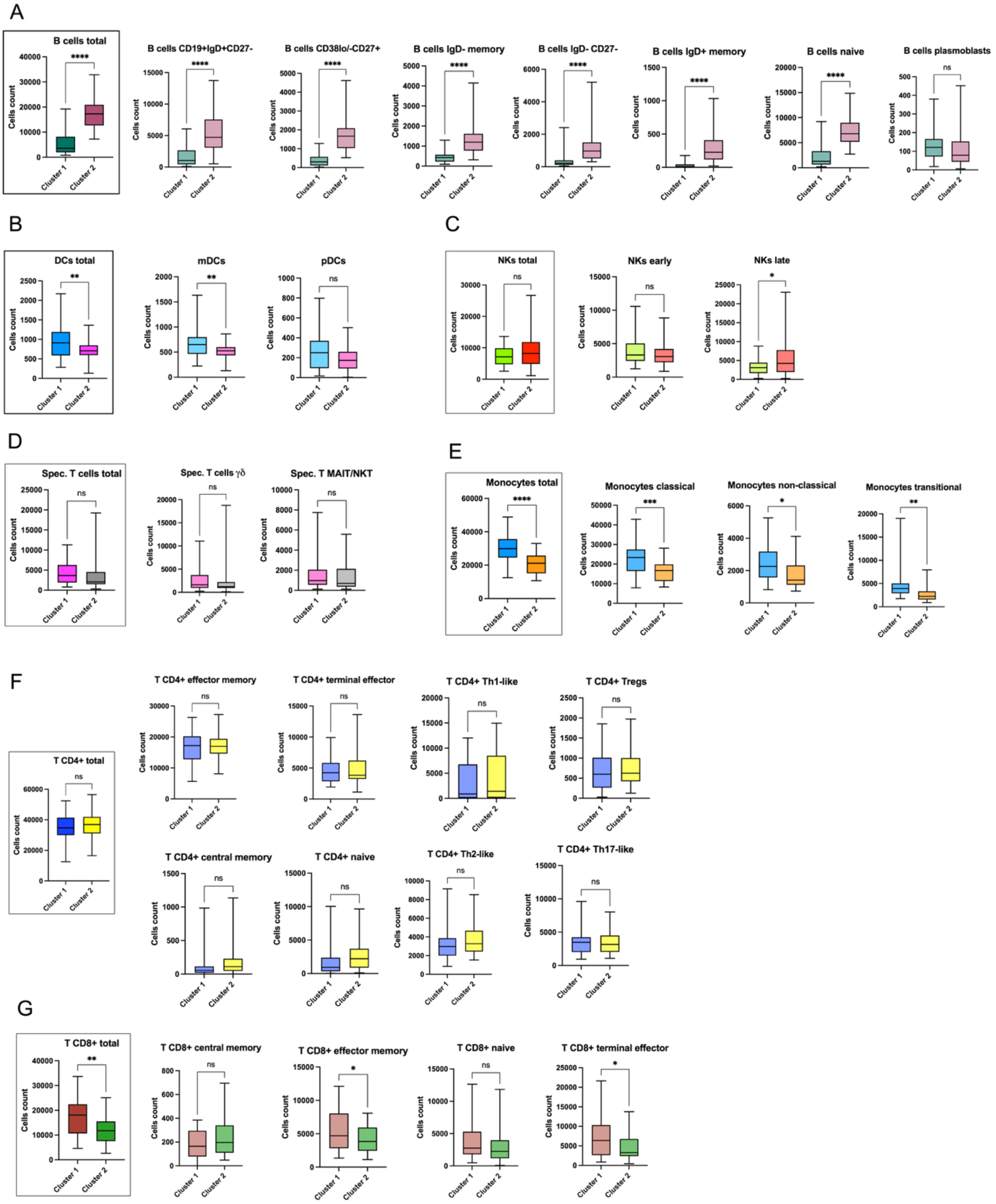


**Figure S2 Differences in immune cell populations** in whole blood between patients assigned to cluster 1 and cluster 2. Graphs in brackets represent major cell types, while the remaining panels show corresponding immune cell subtypes. (A) B cells, (B) dendritic cells (DCs), (C) natural killer cells (NKs), (D) specialized T cells, (E) monocytes, (F) CD4+ T cells, and (G) CD8+ T cells. Statistical significance was assessed using Welch's t-test. * P < 0.05, ** P < 0.005, *** P < 0.0005.

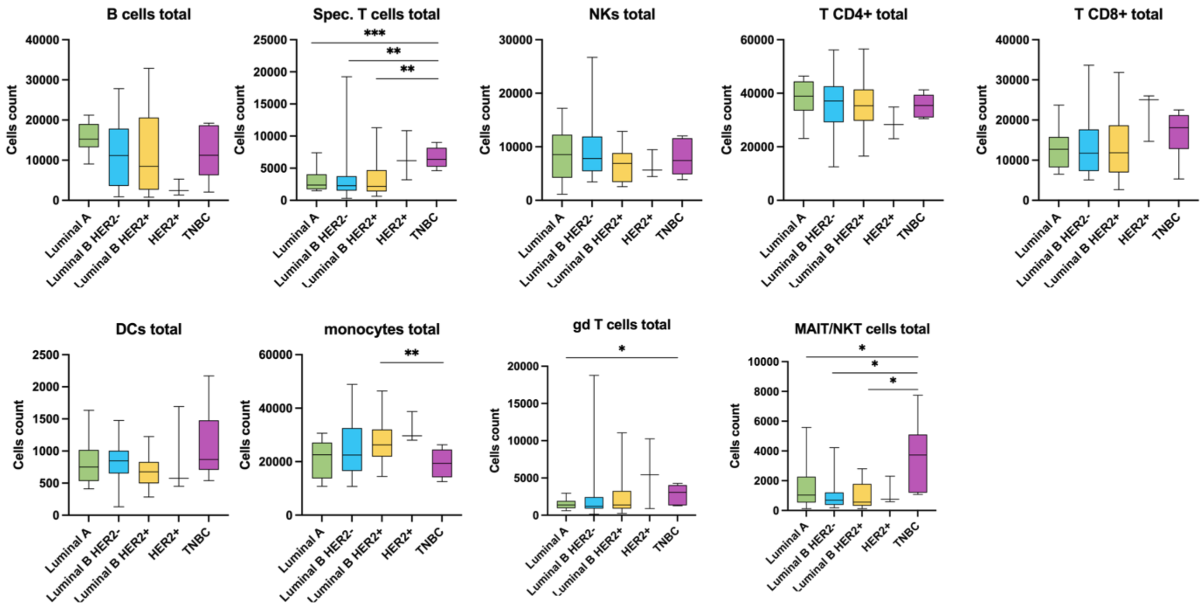


**Figure S3 Differences in immune cell counts** between breast cancer patients stratified by molecular subtype: Luminal A, Luminal B HER2−, Luminal B HER2+, HER2+ non-luminal, and triple-negative breast cancer (TNBC). Statistical significance was assessed using Welch's t-test. * $P < 0.05$, ** $P < 0.005$, *** $P < 0.0005$.

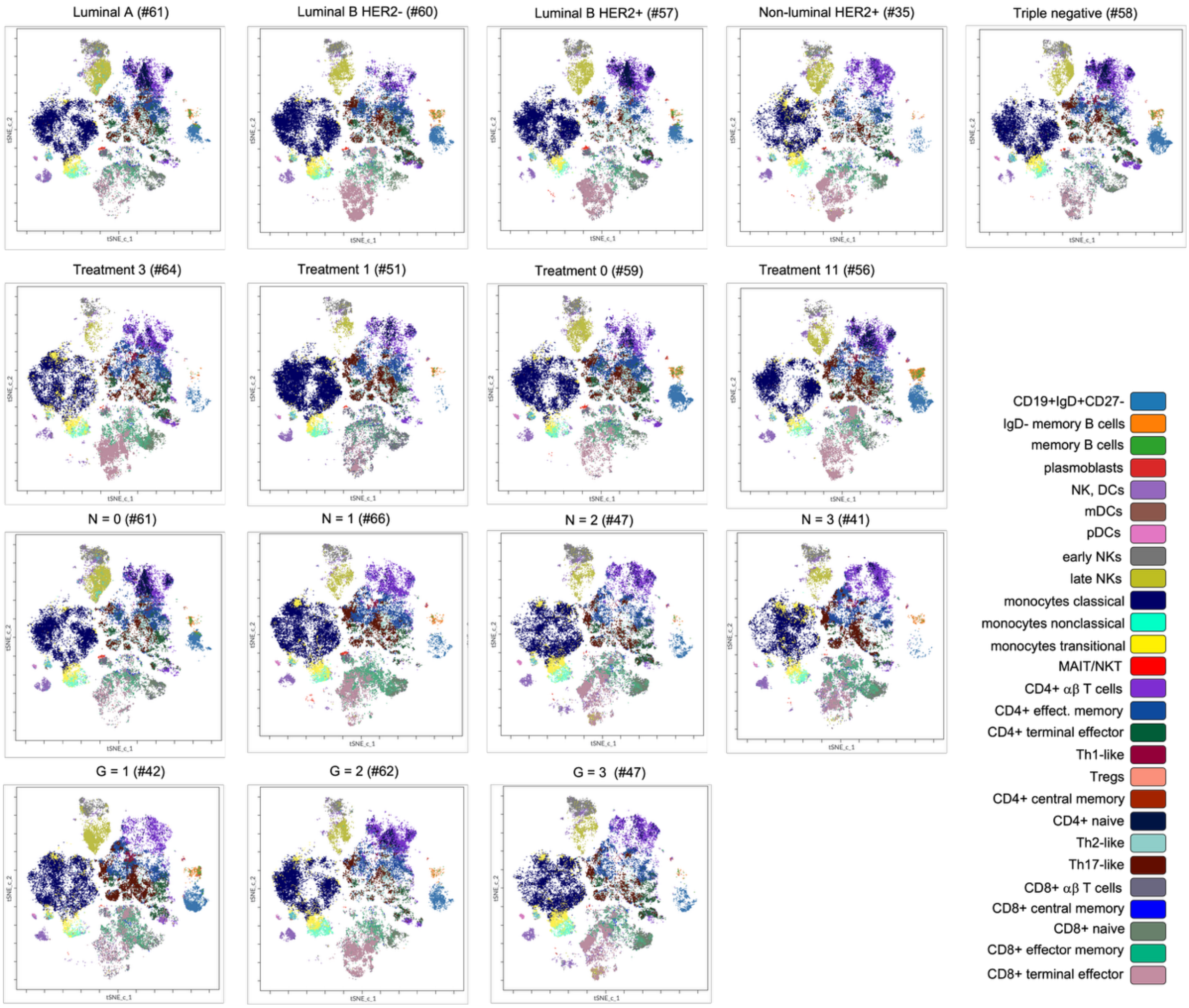


**Figure S4 viSNE maps** generated for individual patients based on various clinical criteria, including molecular subtype, treatment status, nodal status, and tumor grade.

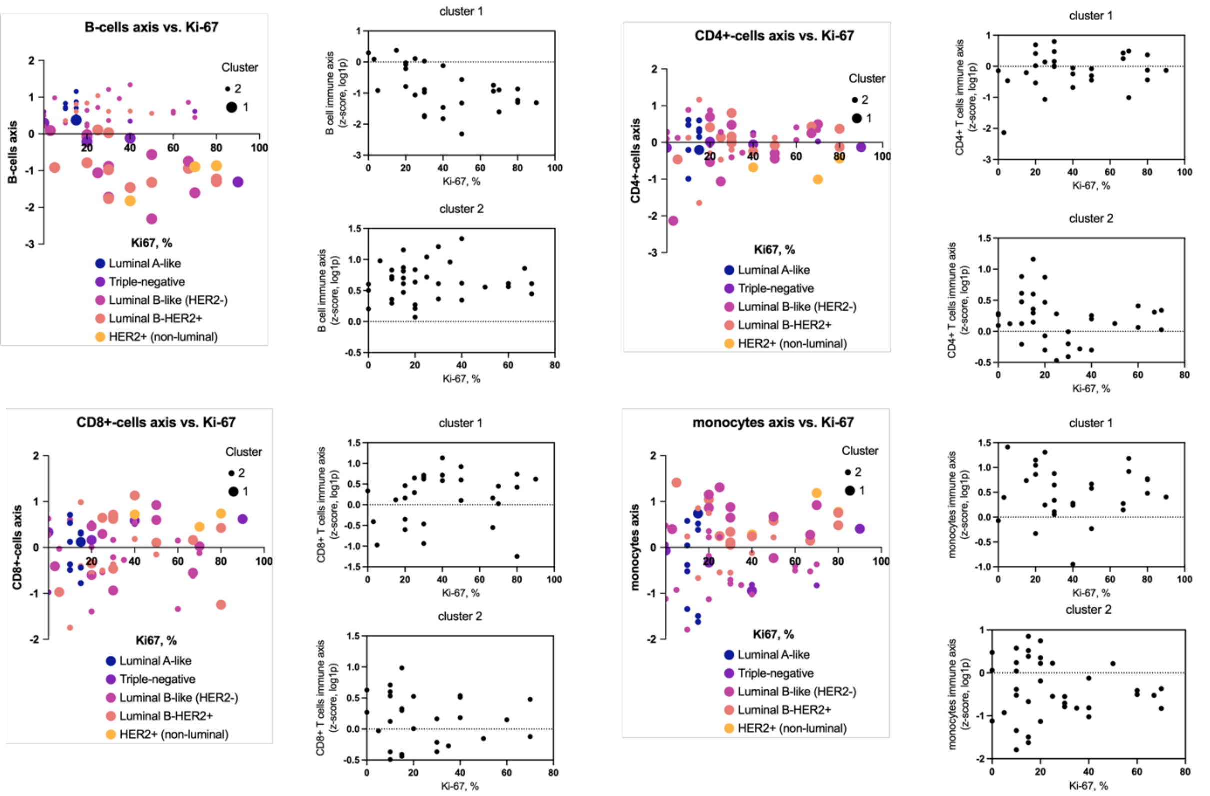


**Figure S5 Relationship between selected immune cell populations** (B cells, CD4+ cells, CD8+ cells and monocytes) and tumor Ki67 status. The immune axis (y), defined as the z-score of log1p-transformed cell counts is ploted versus histological Ki-67 status of tumor (0-100%). Data is presented aggregated, and also splitted into clusters.

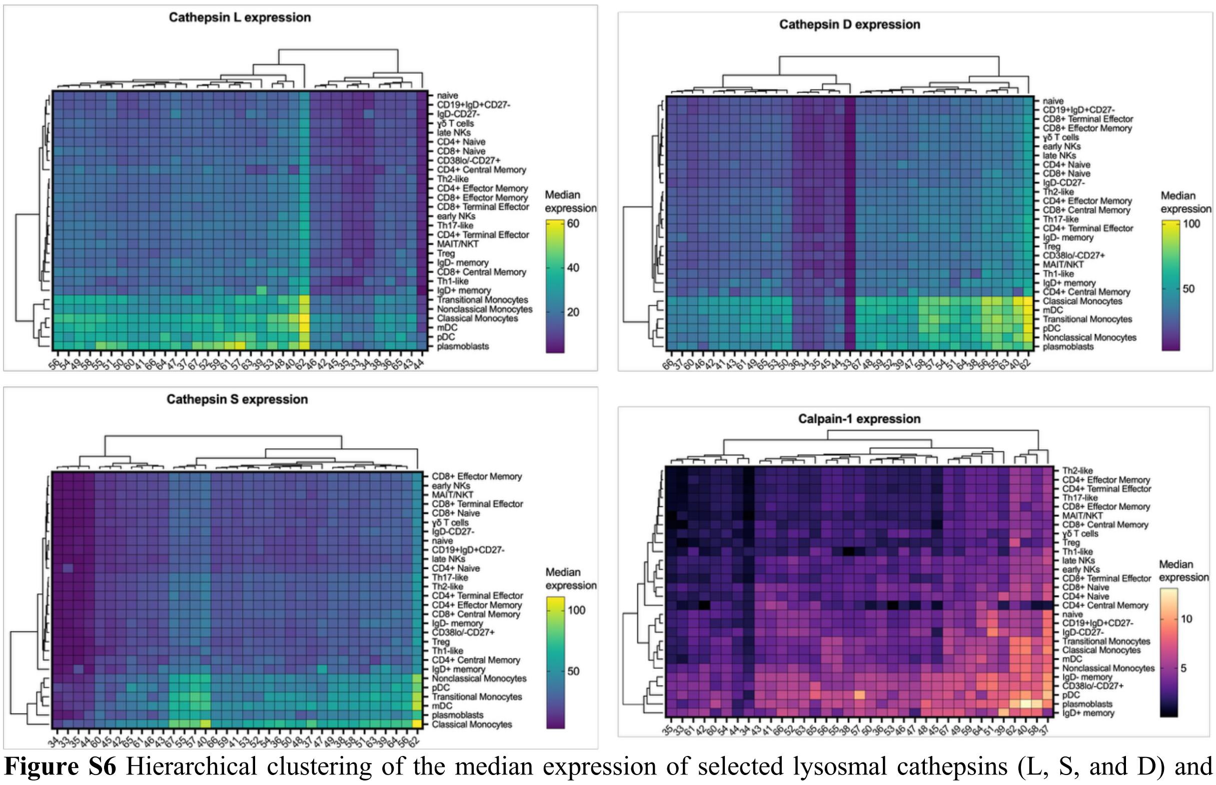


**Figure S6** Hierarchical clustering of the median expression of selected lysosomal cathepsins (L, S, and D) and calpain-1 across individual immune cell types in breast cancer patients.

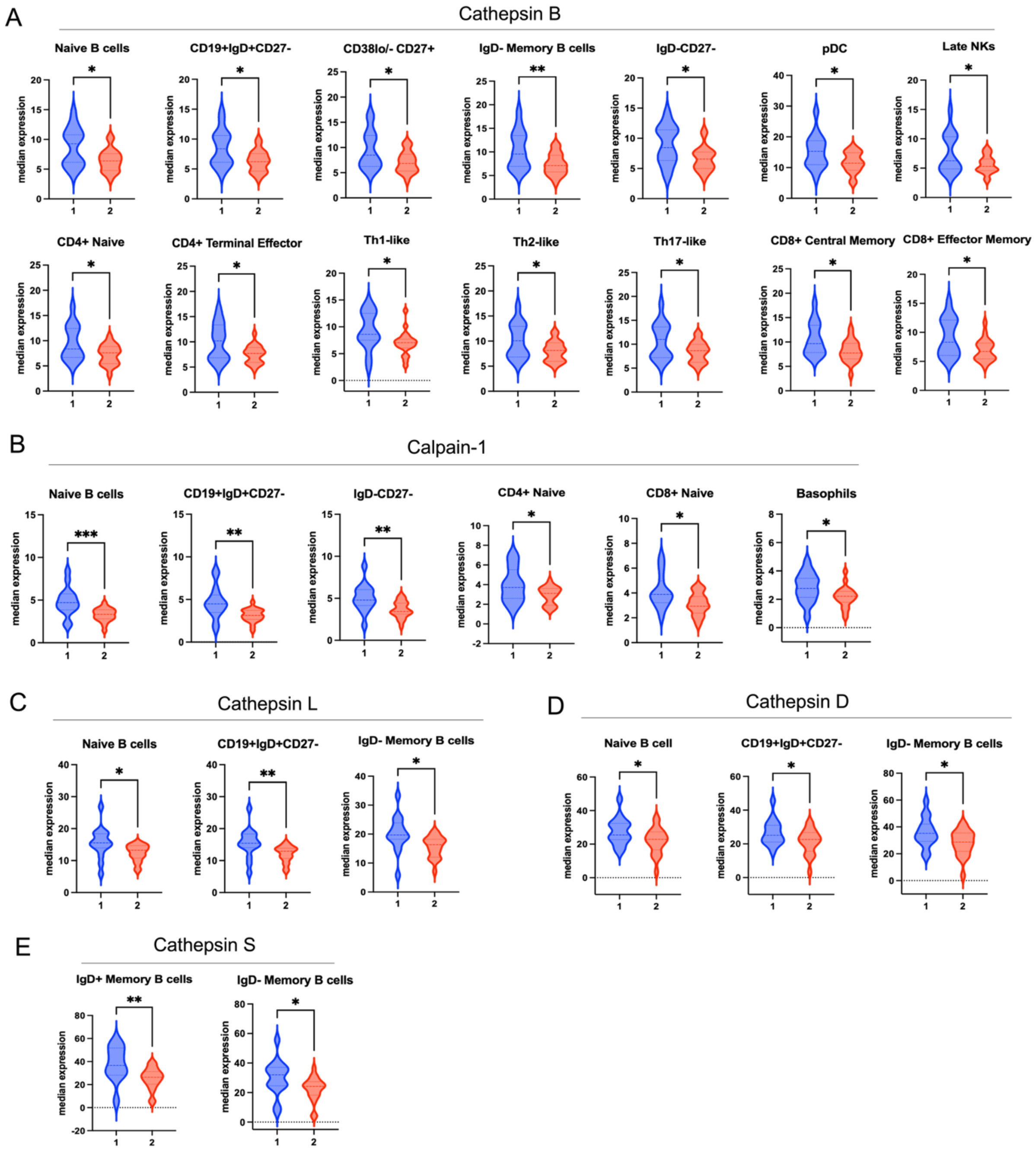


**Figure S7 Differences in median expression levels of selected proteases** in immune cells between patient clusters, including cathepsin B (A), calpain-1 (B), cathepsin L (C), cathepsin D (D), and cathepsin S (E). Only cell populations with statistically significant differences are shown. Statistical significance was determined using Welch's t-test. * $P < 0.05$, ** $P < 0.005$, *** $P < 0.0005$.

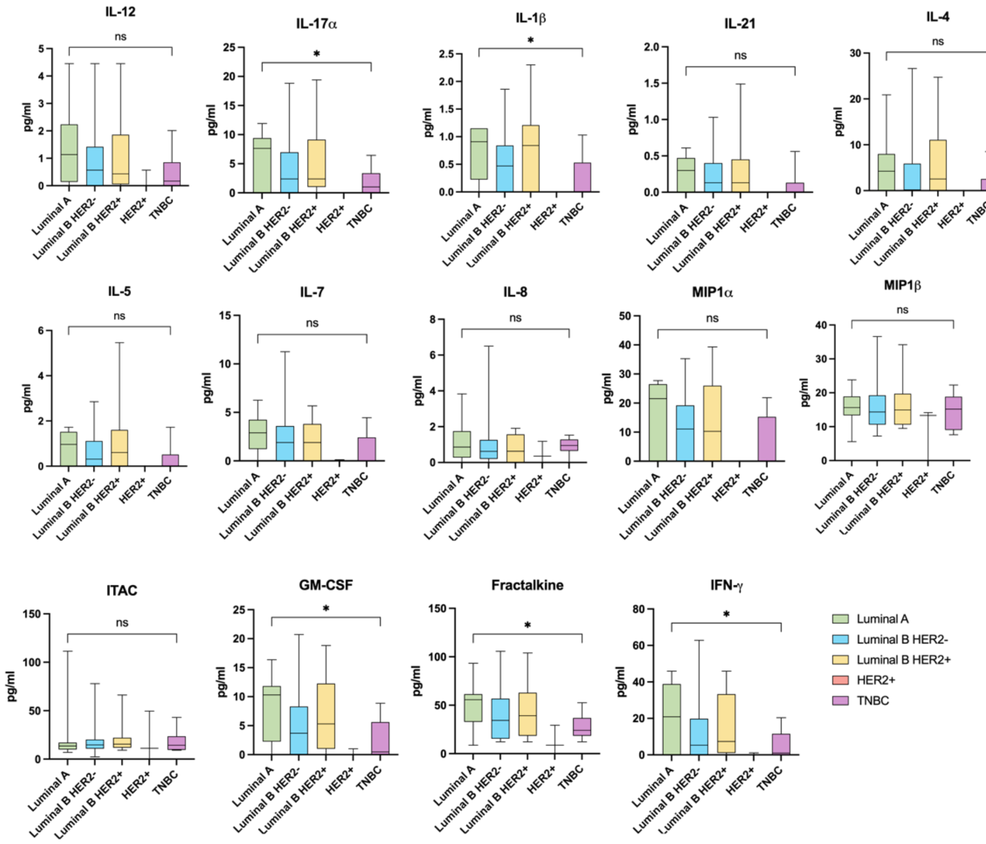


**Figure S8 Serum concentrations of cytokines and interleukins** in breast cancer patients stratified by molecular subtype (Luminal A, Luminal B HER2-, Luminal B HER2+, HER2+ non-luminal, and triple-negative breast cancer). Data are presented as box plots. Statistical significance was determined using Welch's t-test. * $P < 0.05$, ** $P < 0.005$, *** $P < 0.0005$.

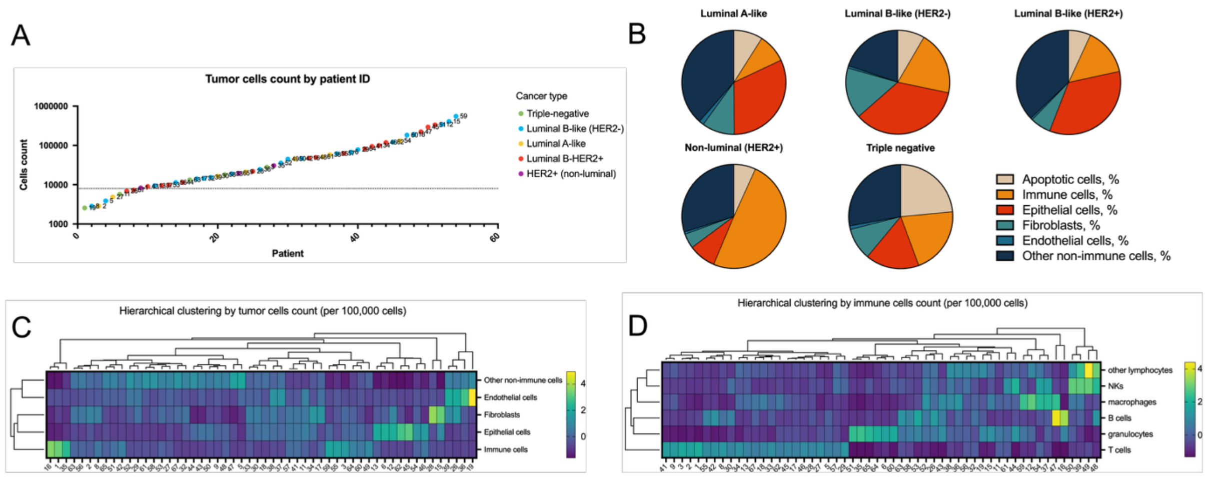


**Figure S9 Analysis of tumor cells in breast cancer samples.** (A) Number of cells collected from individual tumor samples. (B) Pie charts showing the relative proportions of major cell populations, including apoptotic cells, immune cells, epithelial cells, fibroblasts, endothelial cells, and other non-immune cells, across five clinical breast cancer subtypes: luminal A, luminal B HER2−, luminal B HER2+, HER2+ non-luminal, and triple-negative breast cancer. (C,D) Hierarchical clustering of breast cancer patients based on the abundance of major cell populations within tumor samples. (D) Hierarchical clustering of patients based on the abundance of immune cell subsets within tumor samples.

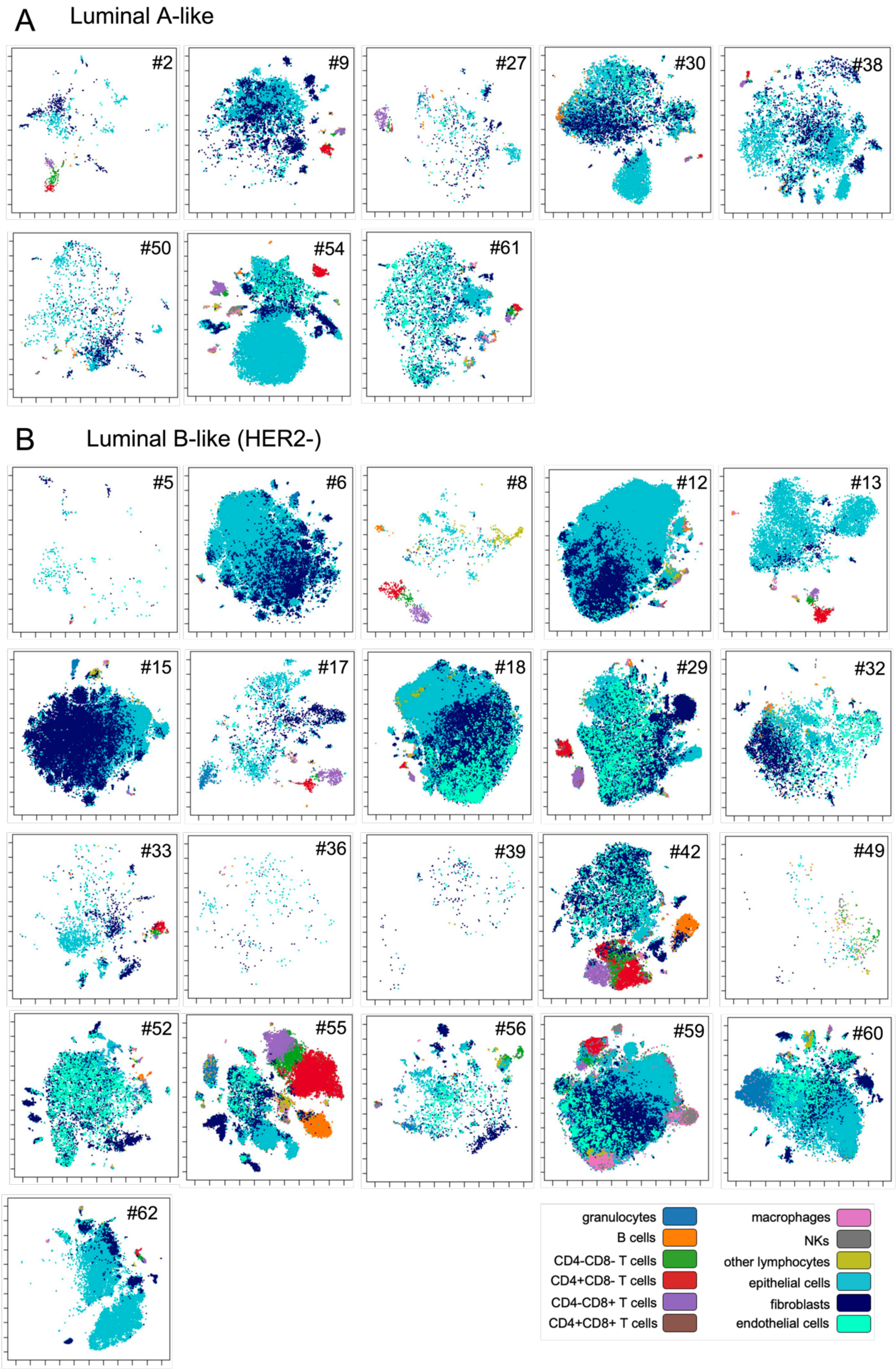


**Figure S10 viSNE maps of individual tumor samples illustrating the single-cell landscape.** Representative maps are shown for patients clinically classified as (A) luminal A and (B) luminal B-like (HER2-).

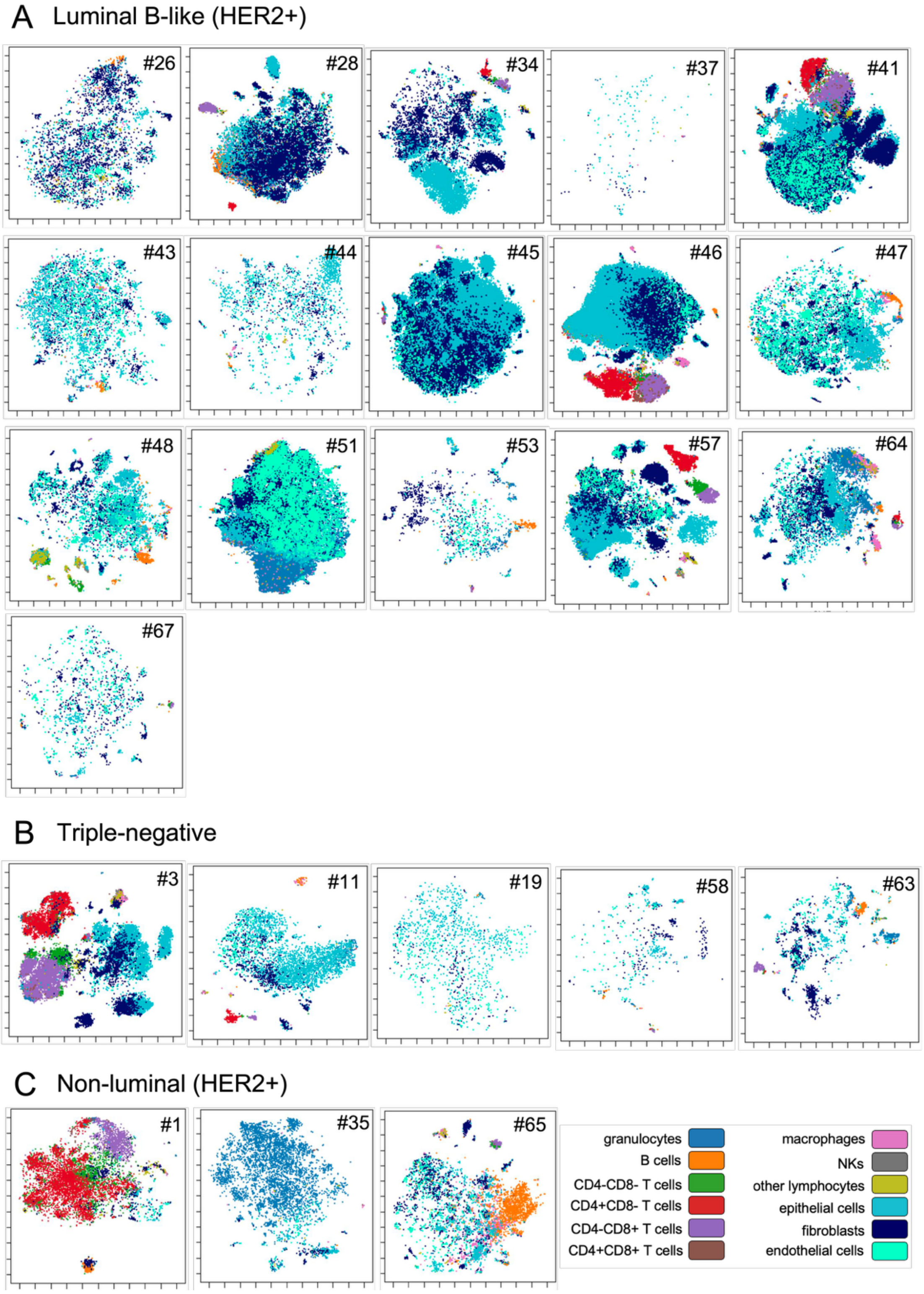


**Figure S11 viSNE maps of individual tumor samples illustrating the single-cell landscape.** Representative maps are shown for patients clinically classified as (A) luminal B-like (HER2+), (B) triple negative and (C) non-luminal (HER2+).

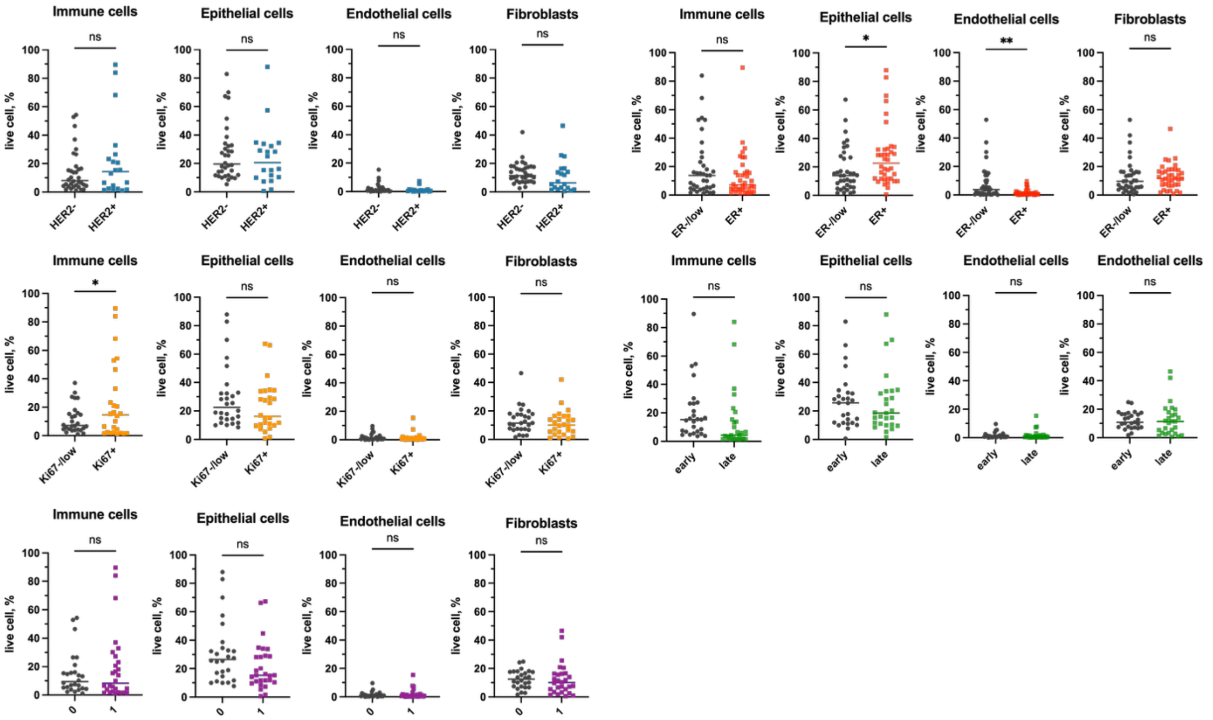


**Figure S12 Differences in the abundance of immune cells, epithelial cells, endothelial cells, and fibroblasts** in tumor samples from patients stratified according to HER2 status (blue), ER status (red), Ki67 expression (orange), tumor stage (green), and neoadjuvant treatment (purple). Statistical significance was assessed using Welch's t-test. * P < 0.05, ** P < 0.005.

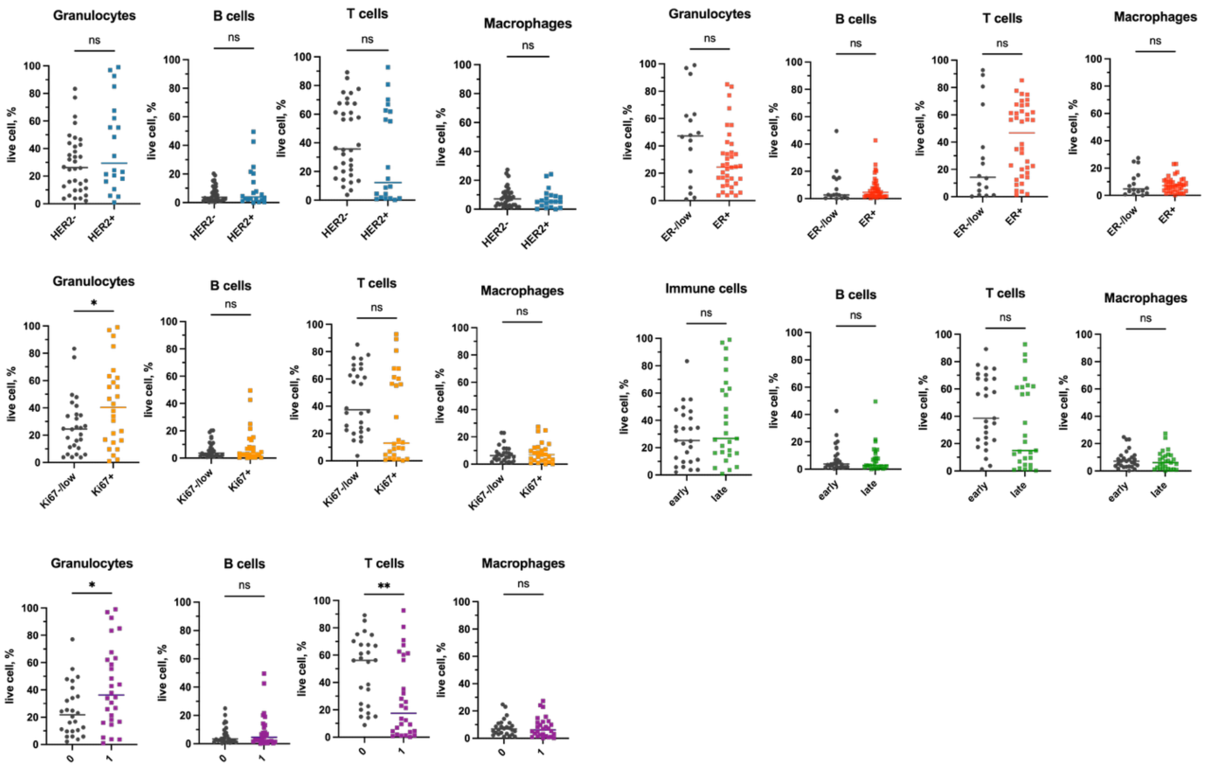


**Figure S13 Differences in the abundance of granulocytes, B cells, T cells, and macrophages** in tumor samples from patients stratified according to HER2 status (blue), ER status (red), Ki67 expression (orange), tumor stage (green), and neoadjuvant treatment (purple). Statistical significance was assessed using Welch's t-test. * P < 0.05, ** P < 0.005.

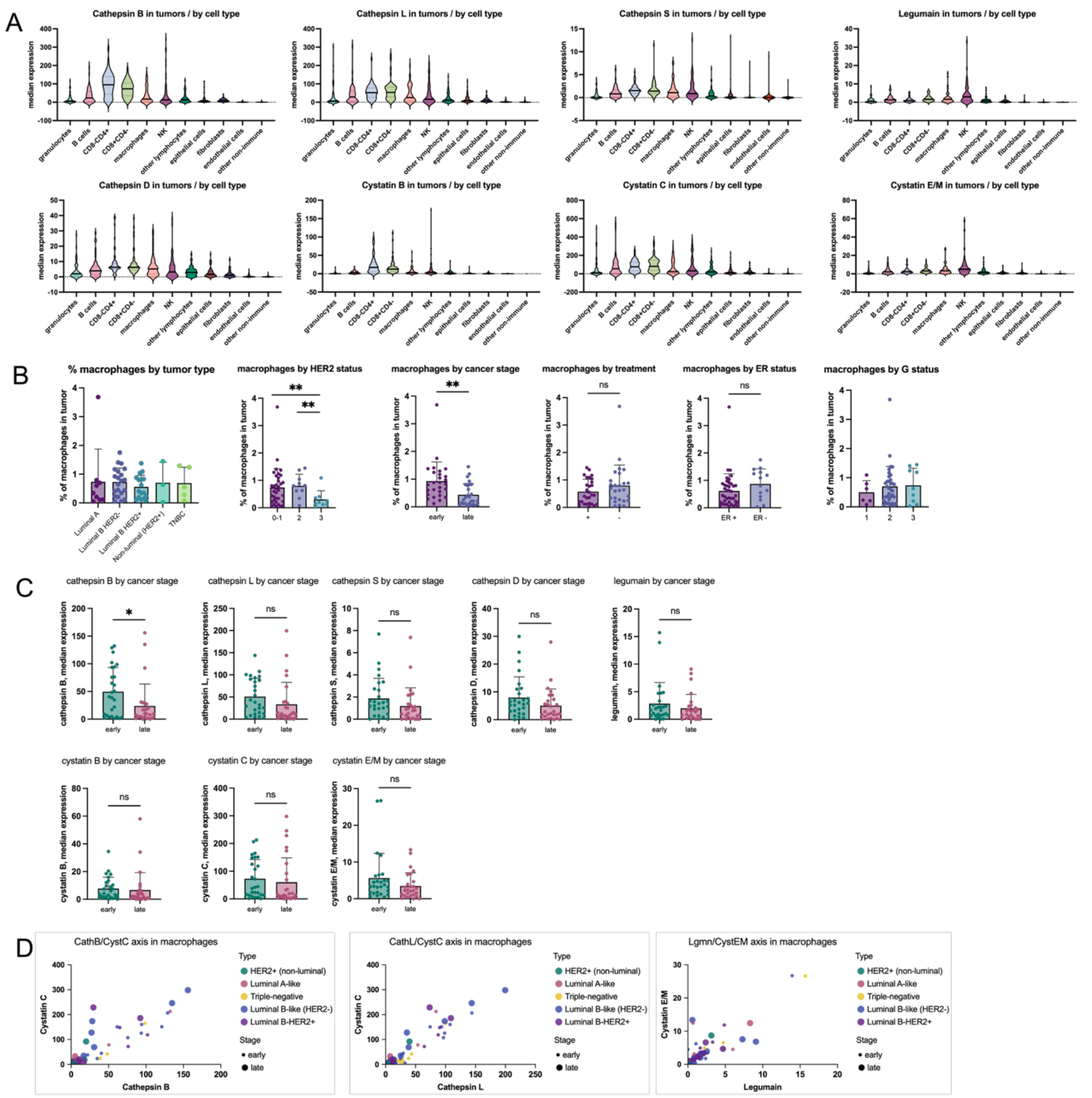


**Figure S14 Analysis of protease and inhibitor expression in breast cancer tumor samples.** (A) Violin plots showing the median expression of lysosomal proteases (cathepsin B, L, S, D, and legumain) and their endogenous inhibitors (cystatins B, C, and E/M) across immune and non-immune cell populations within tumor samples. (B) Percentage of macrophages in tumor samples from patients stratified by clinical breast cancer subtype, HER2 status (0-1, 2, 3), tumor stage (early, late), neoadjuvant treatment (+/-), ER status (ER+, ER-), and tumor grade (1, 2, 3). (C) Differences in the expression of proteases and their inhibitors in macrophages between early- and late-stage tumors. (D) Correlation plots showing the relationships between cathepsin B and cystatin C, cathepsin L and cystatin C, and legumain and cystatin E/M in macrophages, annotated by breast cancer subtype and tumor stage.

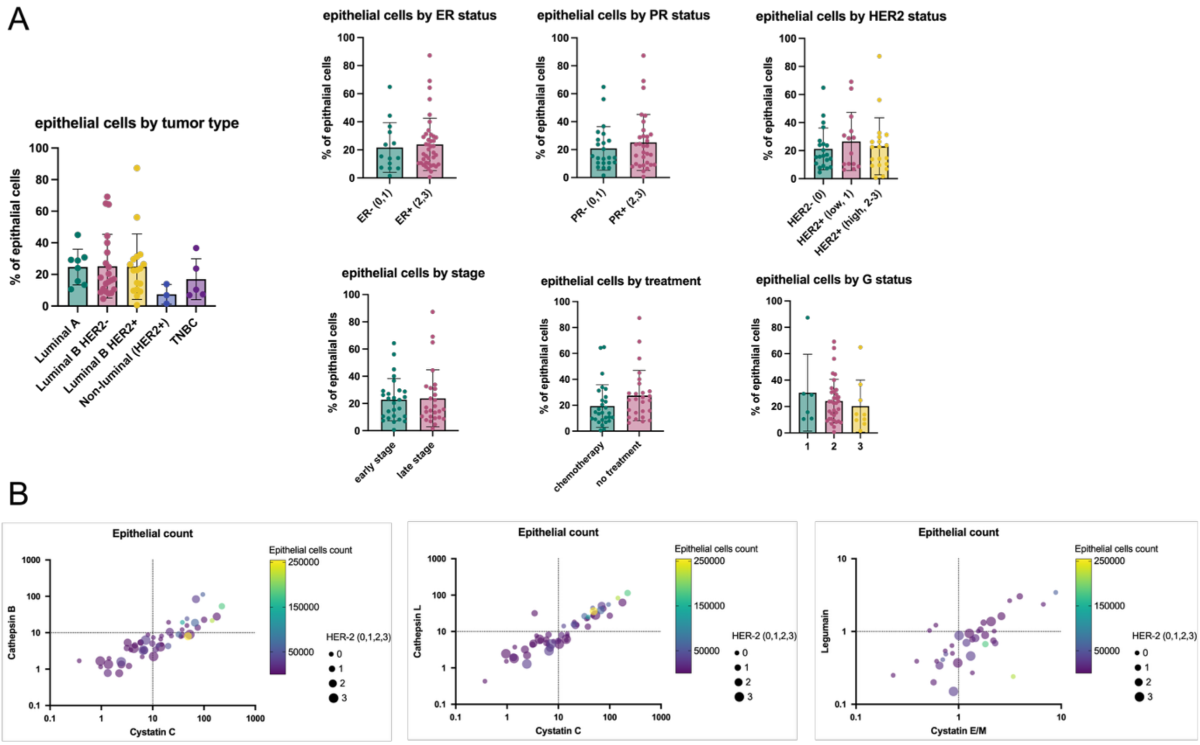


**Figure S15  Analysis of epithelial cells in breast cancer samples.** (A) Differences in the percentage of epithelial cells within tumor samples according to breast cancer subtype, ER status, PR status, HER2 status, tumor stage, neoadjuvant treatment, and tumor grade. (B) Correlation plots showing the relationships between cystatin C and cathepsin B, cystatin C and cathepsin L, and cystatin E/M and legumain in epithelial cells, annotated by epithelial cell abundance and HER2 status.

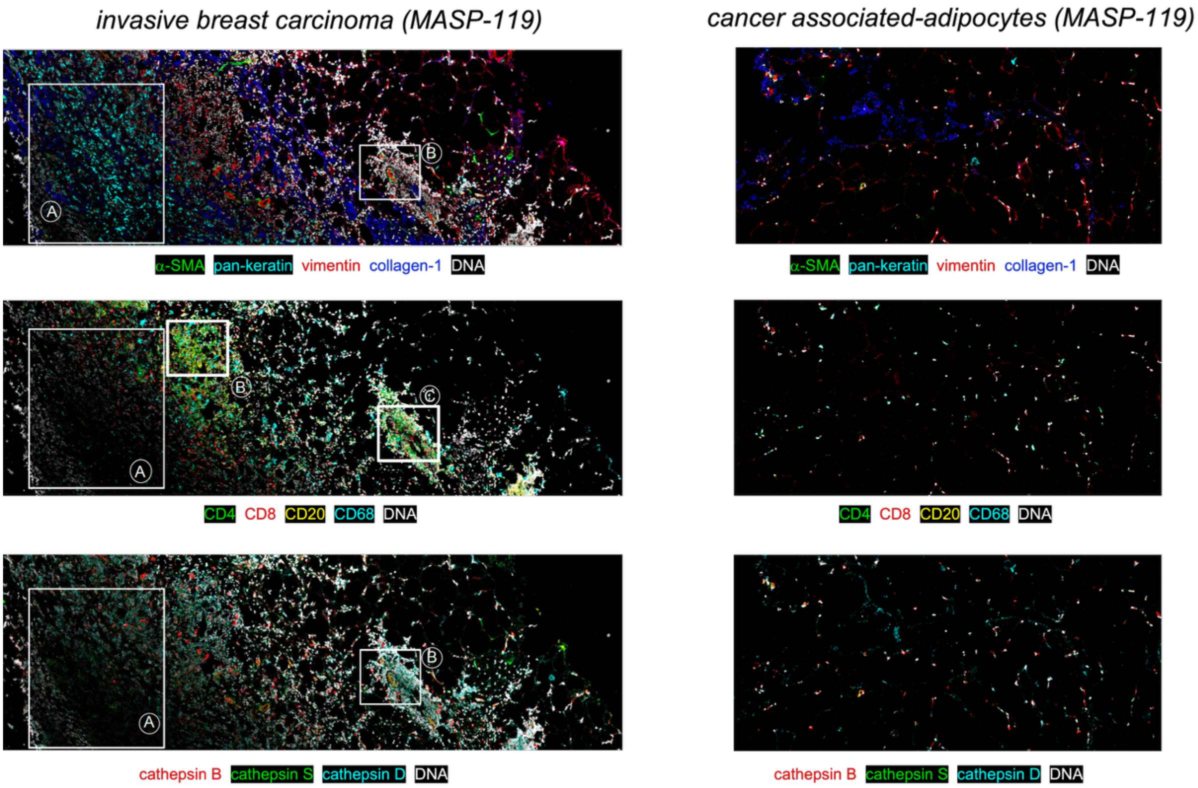


**Figure S16 Spatial analysis of protease expression in breast carcinoma and cancer-associated adipocytes.** The architecture of a representative invasive breast carcinoma and the corresponding cancer-associated adipocytes was visualized based on the spatial expression of structural markers (α-SMA, pan-keratin, vimentin, and collagen I) and infiltrating immune cells (CD4, CD8, CD20, and CD68). In addition, the samples were stained with antibodies against cathepsin B, cathepsin S, and cathepsin D. DNA is shown in white. Each marker was individually scaled to facilitate visualization.

| Target | antibody | clone | vendor | metal |
|---|---|---|---|---|
| Fibroblasts | Anti-FAP | F11-24 | ThermoFisher | 156Gd |
| Fibroblasts | Anti-SMA | 1A4 | ThermoFisher | 175Lu |
| Epithelial cells | Anti-EpCAM (CD326) | 9C4 | BioLegend | 176Yb |
| Epithelial cells | Anti-Cadherin 3 | 67A4 | BioLegend | 141Pr |
| Endothelial cells | Anti-CD31 | WM59 | BioLegend | 171Yb |
| Apoptotic cells | Anti-cleaved Casp-3 (Asp175) | Polyclonal | Cell Signaling | 161Dy |
| Apoptotic cells | Anti-cleaved PARP (Asp214) | Polyclonal | Cell Signaling | 160Gd |
| Lysosome | Anti-LAMP-1 | H4A3 | BioLegend | 142Nd |
| Cathepsin B | Anti-cathepsin B | #173317 | R&D Systems | 162Dy |
| Cathepsin L | Anti-cathepsin L | #204101 | R&D Systems | 173Yb |
| Cathepsin D | Anti-cathepsin D | #185111 | R&D Systems | 163Dy |
| Cathepsin S | Anti-cathepsin S | #248718 | R&D Systems | 113Cd |
| Legumain | Anti-legumain | #312114 | R&D Systems | 174Yb |
| Cystatin C | Anti-cystatin C | #197807 | R&D Systems | 166Er |
| Cystatin B | Anti-cystatin B | #225228 | R&D Systems | 168Er |
| Cystatin E/M | Anti-cystatin E/M | #211515 | R&D Systems | 110Cd |
| Total immne cells | Anti-CD45 | HI30 | BioLegend | 147Sm |
| CD3+ Lymphocytes | Anti-CD3 | UCHT1 | BioLegend | 151Eu |
| CD4+ T cells | Anti-CD4 | RPA-T4 | BioLegend | 155Gd |
| CD8+ T cells | Anti-CD8 | RPA-T8 | BioLegend | 114Cd |
| B cells | Anti-CD20 | 2H7 | BioLegend | 116Cd |
| B cells | Anti-CD19 | HIB19 | BioLegend | 165Ho |
| NK cells | Anti-CD56 | HCD56 | BioLegend | 150Nd |
| Myeloid cells | Anti-CD33 | WM53 | BioLegend | 146Nd |
| Monocytes | Anti-CD16 | 3G8 | BioLegend | 152Sm |
| Monocytes | Anti-CD14 | M5E2 | BioLegend | 153Eu |
| Dendritic cells | Anti-CD11c | c 3.9 | BioLegend | 143Nd |
| Macrophages | Anti-CD68 | Y1/82A | BioLegend | 144Nd |
| Eosinophils | Anti-CD15 | W6D3 | BioLegend | 164Dy |
| T regs | Anti-CD25 | BC96 | BioLegend | 169Tm |
| Neutrophils | Anti-CD66b | 6/40c | BioLegend | 209Bi |

**Table S1 Metal-conjugated antibody panel for mass cytometry profiling of breast tumor architecture.**
